# Defining and Engaging a Novel rTMS Target for Nicotine Craving in Psychotic Disorders

**DOI:** 10.1101/2025.09.24.25334160

**Authors:** Heather Burrell Ward, Sophia H. Blyth, Simon Vandekar, Baxter P. Rogers, Gulcan Yildiz, Jillian G. Connolly, Brett Clementz, Elliot Gershon, Matcheri Keshavan, Shashwath Meda, Godfrey Pearlson, Carol Tamminga, Mark A. Halko, Roscoe O. Brady

## Abstract

Tobacco use is the top preventable cause of early mortality in schizophrenia, but the underlying pathophysiology remains unknown. In schizophrenia, small studies have linked default mode network (DMN) organization to tobacco use and showed that nicotine normalizes DMN disorganization. We sought to 1) validate the relationship between DMN organization and tobacco use using a large psychosis-spectrum sample (Bipolar-Schizophrenia Network on Intermediate Phenotypes 2, B-SNIP2); and 2) test if targeting this network with single and multiple sessions of repetitive transcranial magnetic stimulation (rTMS) affects craving. In B-SNIP2, we tested associations between DMN connectivity and tobacco use. In the Single Session DMN-targeted rTMS study, individuals received single rTMS sessions (intermittent theta burst stimulation, iTBS; continuous theta burst stimulation, cTBS; sham) with pre-/post-neuroimaging and craving assessment. In the Accelerated, Multi-Session DMN-targeted cTBS study, individuals received 5 sessions of cTBS with pre-/post-neuroimaging and craving assessment. In B-SNIP2 (n=596), current smokers had lower DMN connectivity than former (p=.017) and never smokers (p=.021). These differences were also observed in the psychosis group (current vs. former p=.044; current vs. never p=.011). In the Single Session DMN-targeted rTMS study (n=10), there was a nonsignificant treatment*time interaction (p=.059) where iTBS increased craving (p_adj_=.015) compared to cTBS and sham. In the Accelerated, Multi-Session DMN-targeted cTBS study (n=12), DMN-targeted cTBS reduced craving after each session (p<.001) and reduced DMN connectivity (p=.052). We identified a mechanism of nicotine use in psychosis and demonstrated that engaging this target reduces craving, suggesting a novel target for nicotine interventions in psychosis.

## Introduction

Tobacco use is the top preventable cause of early mortality in schizophrenia, leading to a 20-year decreased life expectancy compared to the general population (1, 2). This is because the prevalence of tobacco use in schizophrenia is upwards of 60%, three-times the prevalence in the general population. Moreover, current smoking cessation treatments are significantly less effective for people with schizophrenia (3).

Even after decades of research, the underlying pathophysiology to explain the high prevalence of tobacco use in schizophrenia remains unknown. To date, investigators have largely focused on differences in reward circuitry between schizophrenia and healthy smoking populations (4, 5). Unfortunately, this has not led to more effective smoking cessation treatments in schizophrenia. What if, in addition to reward circuits well-established in addiction, there was another brain circuit driving nicotine use in schizophrenia? If we could treat this schizophrenia-specific circuit pathology, could we develop more effective smoking cessation treatments for this population?

To look beyond reward circuitry, in previous work, we used an agnostic, data-driven connectome-wide analysis that identified novel schizophrenia-specific brain circuits linked to nicotine use (6). We observed that default mode network (DMN) organization was linked to daily cigarette use in schizophrenia, but not in controls. While promising, this finding was observed in only 35 people and requires replication in a larger, more heterogeneous sample.

Therefore, in this manuscript, we first use a large psychosis-spectrum sample and healthy controls (Bipolar-Schizophrenia Network of Intermediate Phenotypes 2, B-SNIP2, n=596, Figure 1A) to validate the relationship between DMN organization and tobacco use in psychosis. Only in the psychosis group, we observe that 1) current smokers have lower DMN connectivity than former and never smokers and 2) that higher daily cigarette use is associated with lower DMN connectivity.

**Figure 1.**
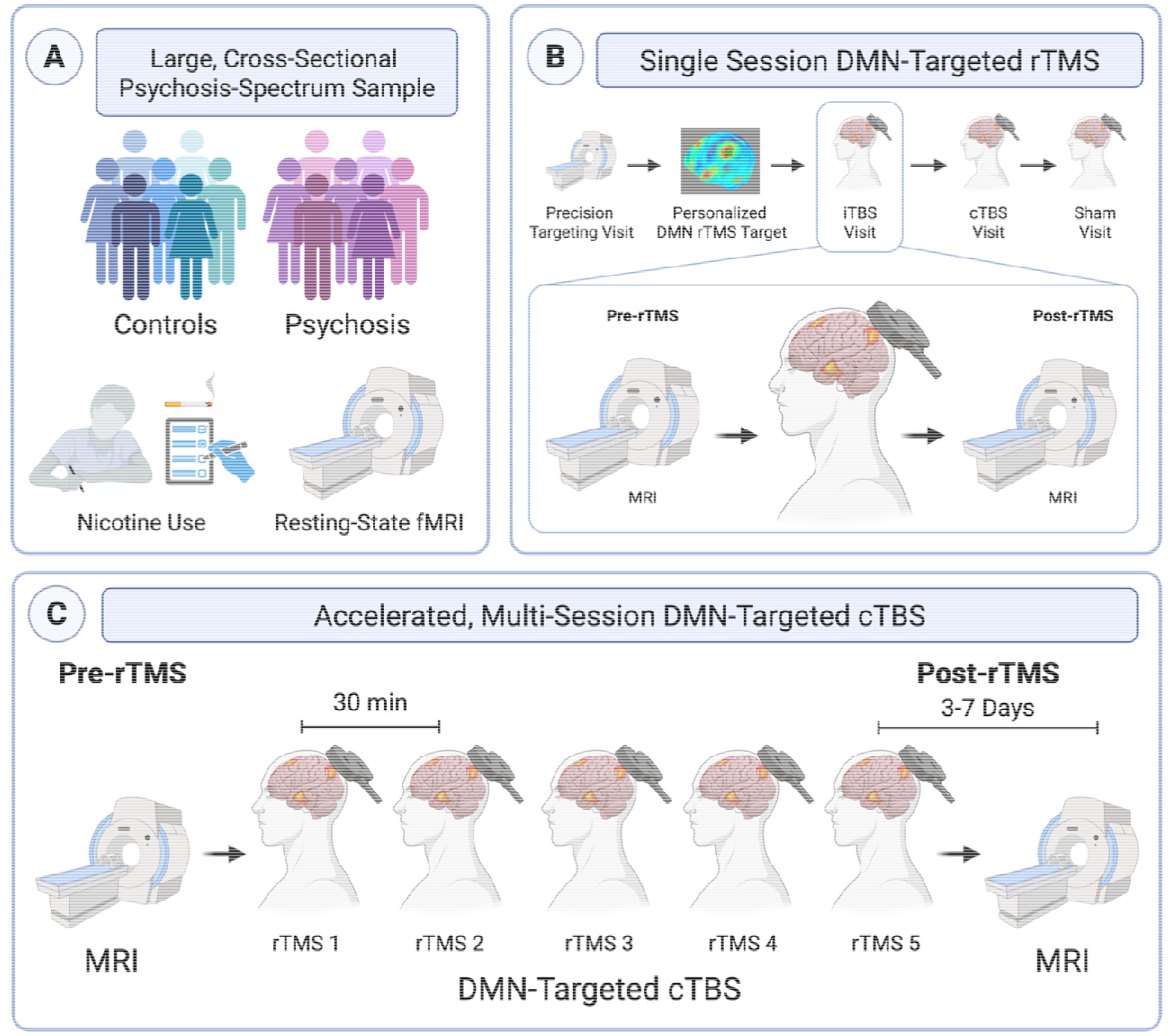
Data Analysis Diagram. In previous work, we used an agnostic, data-driven connectome-wide analysis that identified novel schizophrenia-specific brain circuits linked to nicotine use (6). We observed that the organization of the default mode network (DMN) was linked to daily cigarette use in schizophrenia, but not in controls. In this manuscript, we conducted a series of three experiments to extend and validate this relationship. First, we used large cross-sectional sample of people with psychosis-spectrum disorders and healthy control (Bipolar-Schizophrenia Network of Intermediate Phenotypes 2, B-SNIP2, n=596) to validate a psychosis-specific relationship between DMN organization and tobacco use (Figure 1A). We then tested this network model of nicotine use in psychosis in two mechanistic repetitiv transcranial magnetic stimulation (rTMS) studies. First, we applied single sessions of DMN-targeted iTBS, cTBS, and sham to determine their effects on nicotine craving (Single-Session DMN-Targeted rTMS, Figure 1B). We observed that single sessions of iTBS targeted to the DMN acutely increased craving compared to cTBS and sham. Given the hypothesized opposing effects of iTBS and cTBS, we increased the stimulation intensity of cTBS and delivered multiple sessions in an accelerated design (Accelerated, Multi-Session DMN-Targeted cTBS, Figure 1C). With a higher stimulation intensity, we subsequently observed that cTBS reliably reduced craving both within- and between-participants. We also observed reduced DMN connectivity following multiple sessions of cTBS, thereby demonstrating evidence of target engagement. Created with BioRender.com.

These findings linking current cigarette use with reduced DMN connectivity are consistent with our own previous work showing that acute nicotine administration *normalizes* DMN connectivity – but only in people with psychosis, not in controls (6). These results suggest that modulating DMN connectivity may be a novel intervention for nicotine craving in psychosis.

To test this *network model* of nicotine use in psychosis, we used repetitive transcranial magnetic stimulation (rTMS) interventions targeted to the DMN (7). rTMS can have excitatory or inhibitory effects depending on its stimulation pattern. Intermittent theta-burst stimulation (iTBS) has been associated with excitatory effects (i.e., increased connectivity), while continuous theta-burst stimulation (cTBS) has been associated with inhibitory effects (8), but these effects have been inconsistent. We identified a reliable, personalized DMN target and applied single sessions of iTBS, cTBS, and sham to determine their effects on nicotine craving, a core feature of substance use that is dynamic, susceptible to intervention, and predicts relapse (Single-Session DMN-Targeted rTMS, Figure 1B) (9).

We observed that single sessions of iTBS targeted to the DMN acutely increased craving compared to cTBS and sham. Given the hypothesized opposing effects of iTBS and cTBS, we increased stimulation intensity of cTBS and delivered multiple sessions in an accelerated design (Accelerated, Multi-Session DMN-Targeted cTBS, Figure 1C). With a higher stimulation intensity, we observed that cTBS reliably reduced craving both within each session and between participants. Multiple cTBS sessions reduced DMN connectivity, thereby demonstrating evidence of target engagement.

Our findings provide first evidence for a potential mechanism of nicotine use in psychosis that suggests a novel target for smoking cessation interventions.

## Methods

### B-SNIP2

#### Participants

Data came from B-SNIP2, a multisite, cross-sectional study of individuals with psychotic disorders aged 18-65 who underwent clinical characterization, cognitive assessment, and structural and resting-state functional brain imaging (Supplement). The final sample (Table 1) included 276 healthy and 320 psychosis-spectrum individuals (schizophrenia = 129, schizoaffective disorder = 124, bipolar disorder with psychotic features = 67).

**Table 1.**
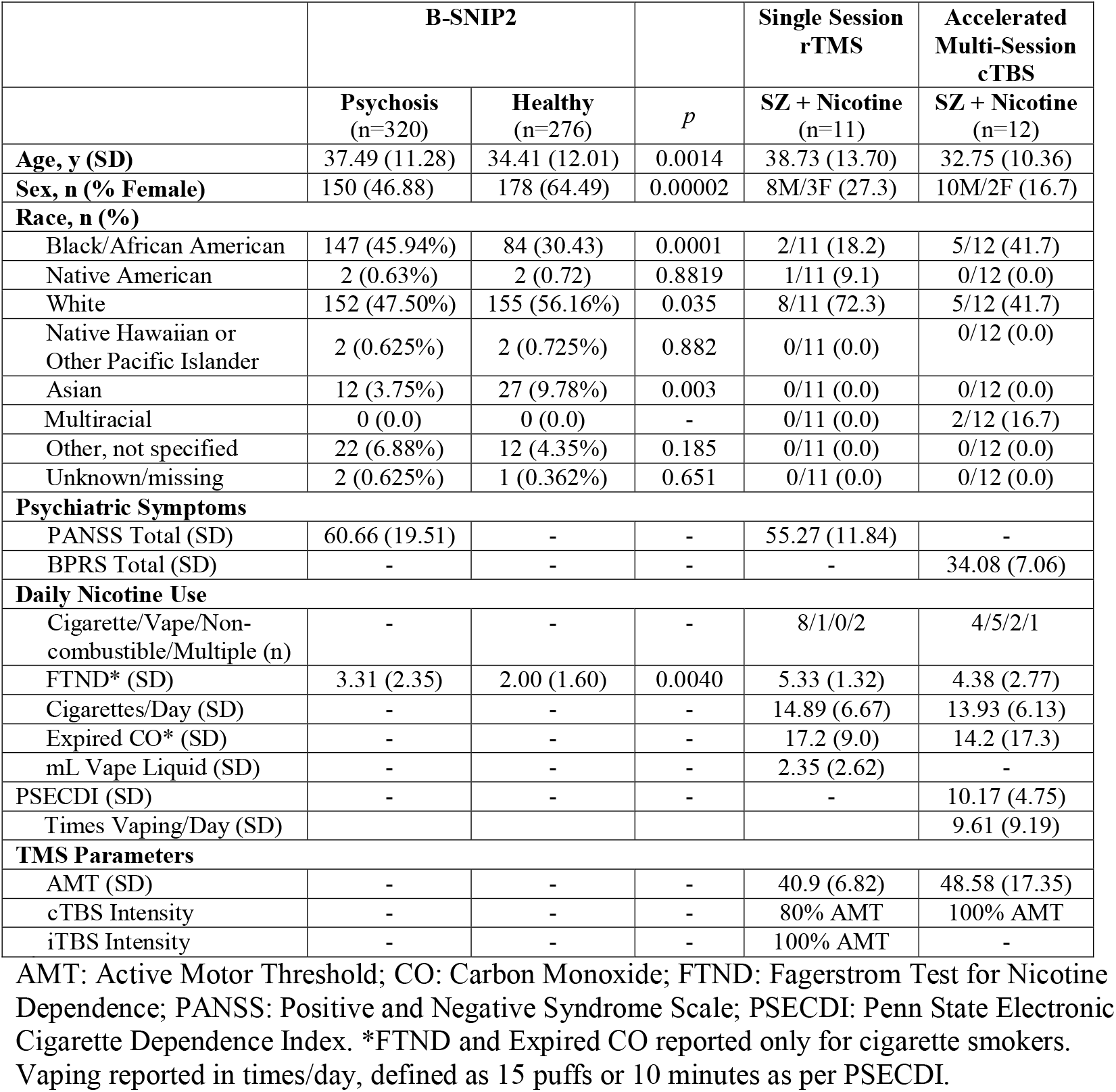
Demographics.

#### Nicotine Assessments

Nicotine dependence was assessed using the Fagerstrom Test for Nicotine Dependence (FTND) (10). Participants were also asked their tobacco use history.

#### MRI Acquisition & Processing

Subjects underwent a T1-weighted structural scan and 8-minute resting-state fMRI scan (rs-fMRI) on a 3T scanner. Preprocessing was performed using the advanced DPARSF module within DPABI V6.0_210501 (Supplemental Table 1). Volume slices were corrected for different signal acquisition times across sites. The time series of images for each subject were realigned. Individual structural images (T1-weighted) were co-registered to the mean functional image after realignment. Transformed structural images were then segmented into gray matter (GM), white matter (WM) and cerebrospinal fluid (CSF) (11). To remove nuisance signals, the Friston 24-parameter model (12) was utilized to regress out head motion effects. To better control for physiological noise, a combined WM/CSF mask was generated from which the top 5 principal components were extracted and signal adjusted using CompCor (13). Linear trends were adjusted since the blood-oxygen-level-dependent (BOLD) signal exhibits low-frequency drifts. The DARTEL tool (14) was used to transform the functional data from individual native space to MNI space. Following this, temporal filtering (0.01–0.08 Hz) was performed. Details in supplement.

### Single-Session DMN-Targeted rTMS in Schizophrenia

#### Participants

Fifteen individuals with schizophrenia or schizoaffective disorder aged 18-65 who currently used nicotine (confirmed for cigarette smokers by expired carbon monoxide ≥ 5ppm) were enrolled in this randomized, sham-controlled, crossover rTMS-fMRI study (Table 1, Figure 1B, Supplemental Figures 1-2, NCT07190352). Details in supplement.

#### Nicotine Assessments

Nicotine use was assessed at every visit. Nicotine dependence was assessed using the FTND (10) and the Penn State Electronic Cigarette Dependence Index (PSECDI) (15). Nicotine craving was assessed two ways: 1) 0-10 visual analogue scale (VAS) immediately before and after each rTMS session; and 2) Tiffany Questionnaire of Smoking Urges (QSU) (16) at the beginning of each visit. Withdrawal was assessed using the Wisconsin Smoking Withdrawal Scale (WSWS, (17)).

#### MRI Acquisition & Processing

Imaging data were collected on a Siemens 3.0-T MRI system (Munich, Germany). Participants each completed 7 MRI scans, including baseline scan and scans immediately before and after each rTMS session (Figure 1B). A total of 270 min of imaging data was collected per participant, including 140 min of resting-state imaging. Briefly, 1-mm^3^ T1-weighted anatomical scans and multiple 10-minute functional runs were acquired (TR 650ms, TE 34.80ms, flip angle 50 degrees, multiband acceleration factor 8, field of view = 207mm, 64 slices, 3-mm^3^ voxels, anterior to posterior phase-encoded). Anatomical images were segmented into GM, WM, and CSF with the Computational Anatomy Toolbox 12 (CAT12, version 12.5; http://www.neuro.uni-jena.de/cat/). Resting-state scans were preprocessed in SPM12 (https://github.com/vuIIS/vuiis-cci-info?tab=readme-ov-file#citing-xnatdax) and were (1) realigned to a mean scan, (2) coregistered with the native space structural scan, then (3) underwent resting-state denoising procedures: bandpass filter (0.01–0.1 Hz), regression of CSF WM, and mean GM signal, regression of 12 motion parameters (6 translation and rotation parameters and their first derivative). All resting-state scans went through a quality assurance procedure that included calculating framewise displacement (FD) and temporal signal to noise ratio (tSNR). Scans with mean FD > 0.5 or a tSNR lower than the 5th percentile of the sample distribution were excluded from analysis. After quality control, there were pre- and post-rTMS scans for 8 cTBS, 9 iTBS, and 6 sham sessions across 9 subjects. Details in supplement.

#### rTMS Protocol

Individuals received single sessions of theta-burst stimulation (Figure 1B & Supplemental Figure 1) applied to an individualized DMN target (see *Individualized DMN Target* below and Supplemental Figure 3) with neuroimaging collected immediately before and after each session. Individuals received one session of iTBS (600 pulses, 100% AMT), cTBS (600 pulses, 80% AMT as per (18)), and sham (coil flipped 180 degrees using 100% AMT iTBS protocol, 600 pulses) on three separate days, separated by at least 2 days to avoid carryover effect (median 6.5 days, mean 13.8 days (SD 22.2), range 2-96 days). rTMS was applied using a MagPro X100 stimulator and an active figure-of-8 coil (Cool B65, MagVenture, Denmark) held tangentially to the scalp with the handle at 45 degrees. rTMS was applied in the standard theta-burst pattern (3 pulses at 50-Hz repeated at a rate of 5-Hz) (8). Order was randomized, and participants were blinded to stimulation type.

### Multi-Session DMN-Targeted cTBS in Schizophrenia

#### Participants

Twelve individuals with schizophrenia or schizoaffective disorder aged 18-65 who currently used nicotine (confirmed by expired carbon monoxide ≥ 5ppm or urine cotinine >200ng/ml) were enrolled in this rTMS-fMRI study (Figure 1C, Table 1, NCT07155096). Details in supplement.

#### Nicotine Assessments

As in the Single-Session rTMS study, nicotine use was assessed at every visit. Nicotine dependence was assessed using the FTND (10) and PSECDI (15). Nicotine craving was assessed two ways: 1) 0-10 VAS immediately before and after each rTMS session and at each MRI visit; and 2) Tiffany QSU at each MRI visit (16). Withdrawal was assessed using the WSWS (17) at each MRI.

#### MRI Acquisition & Processing

Imaging data were collected on 3.0-T Philips Intera Achieva MRI scanner (Philips Healthcare, Andover, MA). Participants completed MRI scans the week before and after the rTMS intervention. As the durability of 5 rTMS sessions is unknown, post-rTMS scans were scheduled 3 to 7 days after rTMS per protocol (mean 4.91 (SD 1.38) days), but due to repeated scheduling conflicts, one participant completed the post-rTMS scan after 12 days. Briefly, 1-mm^3^ T1-weighted anatomical scans and multiple 10-minute functional runs were acquired (TR 2000ms, TE 28.0ms, flip angle 90 degrees, field of view = 240mm, 38 slices, 3-mm^3^ voxels, anterior to posterior phase-encoded). Anatomical images were segmented into GM, WM, and CSF with CAT12 (version 12.5; http://www.neuro.uni-jena.de/cat/). Resting-state scans were preprocessed in SPM12 (https://github.com/vuIIS/vuiis-cci-info?tab=readme-ov-file#citing-xnatdax) and were (1) realigned to a mean scan, (2) coregistered with the native space structural scan, then (3) underwent resting-state denoising procedures: bandpass filter (0.01– 0.1 Hz), regression of CSF, WM, and mean GM signal, regression of 12 motion parameters (6 translation and rotation parameters and their first derivative). All resting-state scans went through a quality assurance procedure that included calculating FD and tSNR. Scans with mean FD > 0.5 or tSNR lower than the 5th percentile of the sample distribution were excluded from analysis. After quality control, there were pre-/post-rTMS scans for 10 participants. Details in supplement.

#### Multi-Session rTMS Protocol

Individuals received 5 sessions of cTBS (600 pulses, 100% AMT, Figure 1C) applied to an individualized DMN target (see *Individualized DMN Target* below) in a one-day accelerated protocol (30-minute inter-session interval) with pre-/post-rTMS neuroimaging. rTMS was applied using a MagPro X100 stimulator and an active figure-of-8 coil (Cool B65, MagVenture, Denmark) held tangentially to the scalp with the handle at 45 degrees. rTMS was applied in the standard theta-burst pattern (3 pulses at 50-Hz repeated at a rate of 5-Hz) (8). Details in supplement.

### Individualized DMN Targeting

To identify an individualized DMN map for rTMS targeting, a standard DMN template (19) was warped into native space and applied to the participant’s pre-rTMS scan. In each participant, the resultant connectivity maps yielded a correlation cluster in the left posterior inferior parietal lobule (IPL, Supplemental Figure 3). A target was placed in the averaged center of the left posterior IPL correlation cluster (formed from the overlay of the left posterior IPL clusters derived from connectivity maps) on the cortical surface using Brainsight neuronavigation software (Rogue Research, Inc.). This individualized rTMS target selected in the left parietal region of the DMN was used as the rTMS target for all sessions in both Single-Session DMN-Targeted rTMS and Accelerated Multi-Session DMN-Targeted cTBS.

### Neuroimaging Analysis - Calculation of Whole-Network Connectivity Values

For the B-SNIP2, Single-Session rTMS, and Accelerated Multi-Session cTBS analyses, we calculated average DMN connectivity as previously by extracting time courses from standard network nodes and averaging connectivity between DMN nodes for each participant (6). We calculated individual values of average DMN functional connectivity by placing 6mm spheres at coordinates corresponding to standard nodes of the DMN (see (20) for coordinates). The time courses of the BOLD signal from the regions of interest (ROIs) were correlated with each other and z-transformed to generate an ROI-to-ROI connectivity matrix. A mean connectivity from this matrix was generated for each participant by averaging connectivity values for the entire DMN. Details in supplement.

### Statistical Analysis

Chi-square tests were used to compare categorical variables and Welch T-tests were used to compare continuous outcomes between groups (Table 1). For B-SNIP2, a linear mixed effects model with robust standard errors was used to model DMN connectivity by age, sex, site, group, tobacco use history, and their interaction.

For Single Session DMN-targeted rTMS, a linear mixed effects model with robust standard errors was used to model craving by treatment time (pre- and post-stimulation), the rTMS stimulation condition (iTBS, cTBS, sham) and their interaction, including a random effect for subject. Tests of treatment effect for each stimulation condition were adjusted across the three groups using Tukey correction.

For Accelerated, Multi-Session DMN-targeted cTBS, we used a linear mixed effects model to fit craving by treatment time and visit number (modeled categorically) including a random intercept for subject. ANOVAs with robust standard errors were used to test the main effect of treatment time across all sessions. Paired t-tests were used to compare connectivity and behavioral measures at pre- and post-rTMS MRI visits. All analyses were conducted in RStudio (Version 2023.03.1+446).

## Results

### B-SNIP2

#### History of Tobacco Use is Related to DMN Connectivity in Psychosis

In B-SNIP2, individuals with psychosis were more likely to currently use tobacco and to have ever-used tobacco (Supplemental Results and Supplemental Figure 4). We observed differences between history of tobacco use and DMN connectivity in the entire sample (F=3.5798, df1=2, df2=583, p=.028, Supplemental Table 2) such that current smokers had lower DMN connectivity than former (Estimate=-0.040, SE=0.017, t(583)=-2.351, p=.019) and never smokers (Estimate=-0.032, SE=0.013, t(583)=-2.31, p=.021). We did not observe a significant interaction between tobacco use history and group (F=0.9256, df1=2, df2=586, p=0.40). In the psychosis group, we observed significant differences in DMN connectivity by tobacco use history (F=3.8003, df1=2, df2=308, Cohen’s d=0.222, p=.023, Figure 2A) such that current smokers had lower DMN connectivity than former smokers (Estimate=-0.046, SE=0.021, t(308)=-2.162, p=.031) and never smokers (Estimate=-0.044, SE=0.018, t(308)=-2.403, d=0.058, p=.017). We did not observe a relationship between DMN connectivity and tobacco use in controls (F=0.2252, df1=2, df2=266, d= p=.799, Figure 2B).

**Figure 2.**
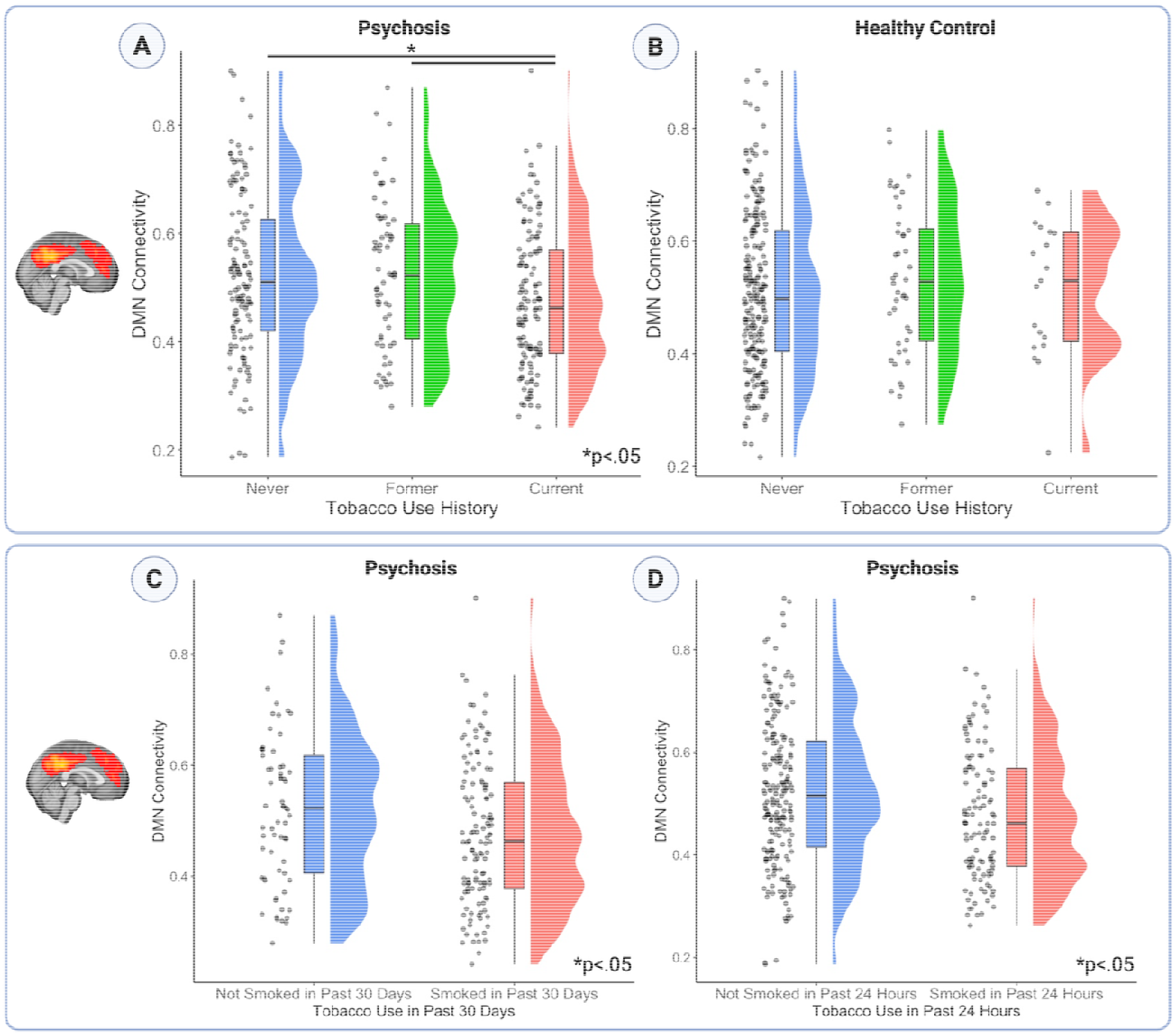
Current Tobacco Use is Related to DMN Connectivity in Psychosis. Using a large, multisite study of individuals with psychotic disorders and controls (B-SNIP2), we observed significant associations between history of tobacco use and DMN connectivity in the psychosi group (p=.023, Figure 2A) but not in controls (p=.66, Figure 2B). In the psychosis group, current smokers had significantly lower DMN connectivity than former (p=.017) and never smoker (p=.021). Moreover, recent tobacco use was associated with lower DMN connectivity in the psychosis group, but not in controls. In the psychosis group, DMN connectivity was significantly lower in individuals who had used tobacco in the last 30 days (0.479 vs. 0.522, p=.038, Figure 2C) and in the last 24 hours (0.478 vs. 0.522, p=.006, Figure 2D) than those who had not used.

#### In Psychosis, Recent Tobacco Use is Associated with Lower DMN Connectivity

Across the entire sample, individuals who used tobacco in the last 30 days and in the last 24 hours had lower DMN connectivity than those who had not (Used in last 30 days: 0.483 vs. Not used: 0.526, T=2.5364, df=227.06, p=.012, CI=[0.0095, 0.076]; Used in last 24 hours: 0.481 vs. Not used: 0.518, T=2.8232, df=220.17, p=.005, CI=[0.011,0.063]). We did not observe a significant interaction between tobacco use history and group (T=0.3817, df=1, p=.5373). In the psychosis group, DMN connectivity was lower in individuals who used tobacco in the last 30 days (Used in last 30 days: 0.479 vs. Not used: 0.522, T=2.0979, df=123.47, Cohen’s d=0.38, p=.038, CI=[0.0024, 0.084]; Figure 2C) and in the last 24 hours (Used in last 24 hours: 0.478 vs. Not used: 0.522, T=2.7879, df=251.9, d=0.35, p=.006, CI=[0.013, 0.075]; Figure 2D). In the control group, we did not observe differences in DMN connectivity based on tobacco use in the past 30 days (0.514 vs. 0.532, T=0.51551, df=37.023, d=0.17, p=.609) or past 24 hours (0.507 vs. 0.516, T=0.26399, df=16.153, d=0.13, p=.795).

#### Higher Cigarette Use is Associated with Lower DMN Connectivity

Daily cigarette use was quantified with the FTND, which provides a binned number of cigarettes smoked per day (≤10; 11-20; 21-30, ≥31). We observed a dose-response relationship where more cigarettes smoked per day was associated with lower DMN connectivity in the entire sample (Spearman, r=-0.21, p=.0095, CI=[-0.37, -0.054]). In a linear model, the interaction between cigarettes per day and group was not a significant predictor of DMN connectivity (Estimate=0.006532, T=0.079, p=.937). In the psychosis group, there was a relationship where more cigarettes smoked per day was associated with lower DMN connectivity (Spearman, r=-0.22, p=.013). This relationship was nonsignificant in controls (Spearman, r=-0.026, p=.915).

### Single-Session DMN-Targeted rTMS in Schizophrenia

#### Single Session DMN-Targeted rTMS Affects Nicotine Craving but not DMN Connectivity

Ten participants with schizophrenia and daily nicotine use completed our randomized, controlled crossover study of individualized, DMN-targeted rTMS (Figure 1B and Supplemental Figures 1-2, Table 1). Participants used various forms of nicotine: cigarettes (9/11), vape (2/11), and non-combustible nicotine (1/11, transdermal patch), were moderately nicotine dependent (mean FTND 5.33, SD 1.32) and smoked on average 14.89 cigarettes/day (SD 6.67). Ten participants completed all study visits and were included in the analysis (Table 1). rTMS was well-tolerated, and there were no serious adverse events. Details in supplement.

There was a nonsignificant treatment by time interaction (X^2^=5.6363, df=2, p=.059) on nicotine craving. Adjusted tests of the treatment effect in each stimulus condition found iTBS significantly increased craving (Estimate=2.5, t=2.521, df=45, p_adj_=.015; Figure 3), with insufficient evidence in cTBS (Estimate=-0.1, t=-0.142, df=45, p_adj_=.88) and sham (Estimate=-0.4, t=-0.371, df=45, p_adj_=.71) conditions. We did not observe a significant treatment by time interaction on average within-DMN connectivity change (X^2^=0.0962, df=2, p=.95), suggesting a single session of rTMS was insufficient to change connectivity of the entire network. There were also nonsignificant effects of treatment (X^2^=1.3409, df=2, p=.51) and time (X^2^=1.8477, df=1, p=.17) on nicotine craving.

**Figure 3.**
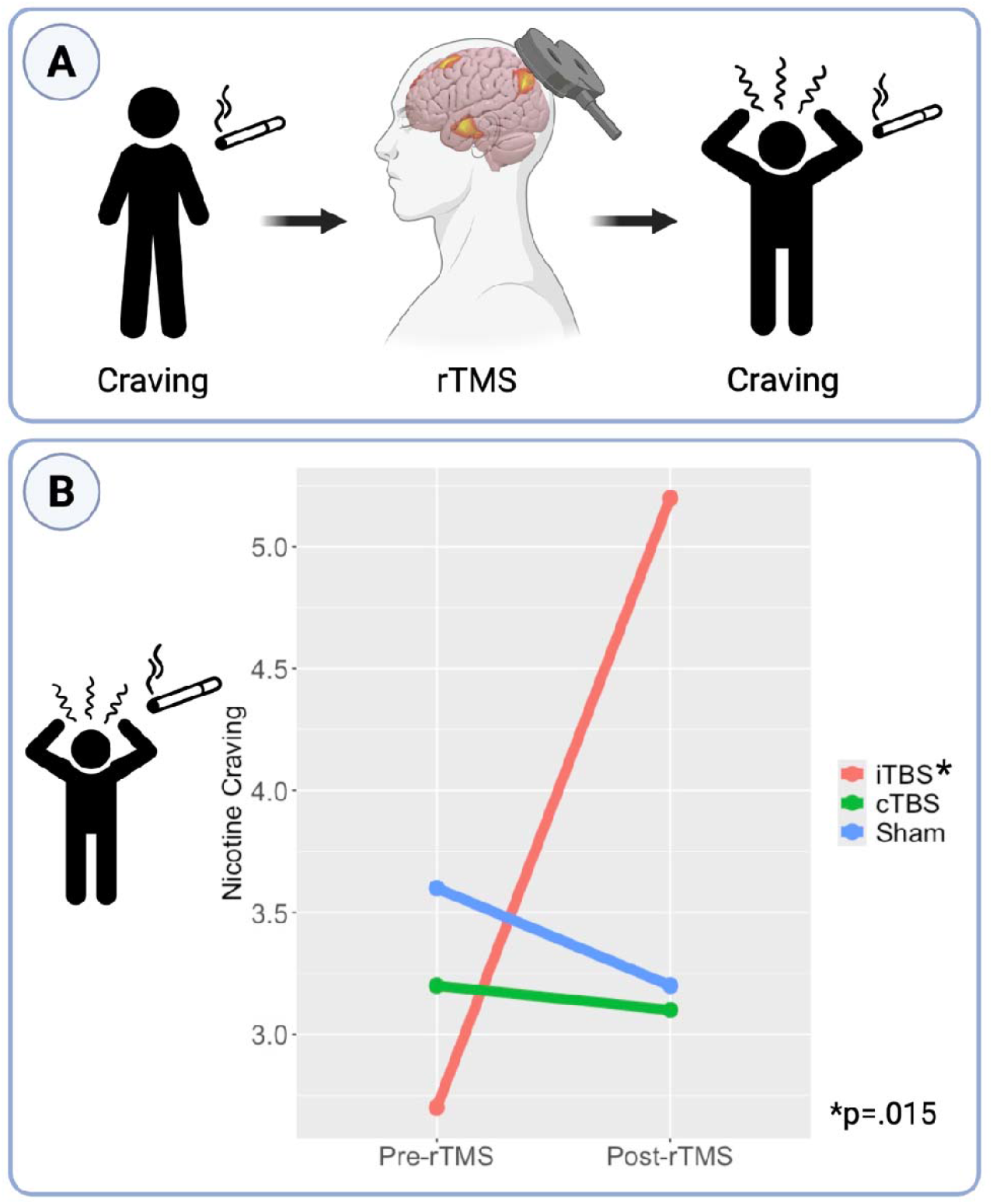
Single Session DMN-Targeted iTBS Increases Nicotine Craving. In a randomized, controlled crossover study of individualized, DMN-targeted rTMS, individuals received a single session of intermittent theta burst stimulation (iTBS), continuous theta burst stimulation (cTBS), and sham stimulation (Figure 3A). Nicotine craving was assessed immediately before and after each single rTMS session using 0-10 visual analog scale. There was a trend-level group x time interaction (p=.059) on nicotine craving such that iTBS significantly increased craving (p_adj_=.015, Figure 3B) compared to cTBS and sham (p>.05). Created with BioRender.com.

### Multi-Session DMN-Targeted cTBS in Schizophrenia

#### Multiple Sessions of DMN-Targeted cTBS Reduce Nicotine Craving

Twelve participants with schizophrenia and daily nicotine use were enrolled in an Accelerated Multi-Session study of individualized, DMN-targeted cTBS (Figure 4A, Table 1, Supplemental Figure 6). Participants used various forms of nicotine: cigarettes (4/12), vape (6/12), and non-combustible nicotine (2/12, transdermal patch, lozenge, or pouch), were moderately nicotine dependent (mean FTND 4.38, SD 2.77) and smoked on average 14.89 cigarettes/day (SD 6.67). rTMS was well-tolerated, and there were no serious adverse events. Details in supplement.

**Figure 4.**
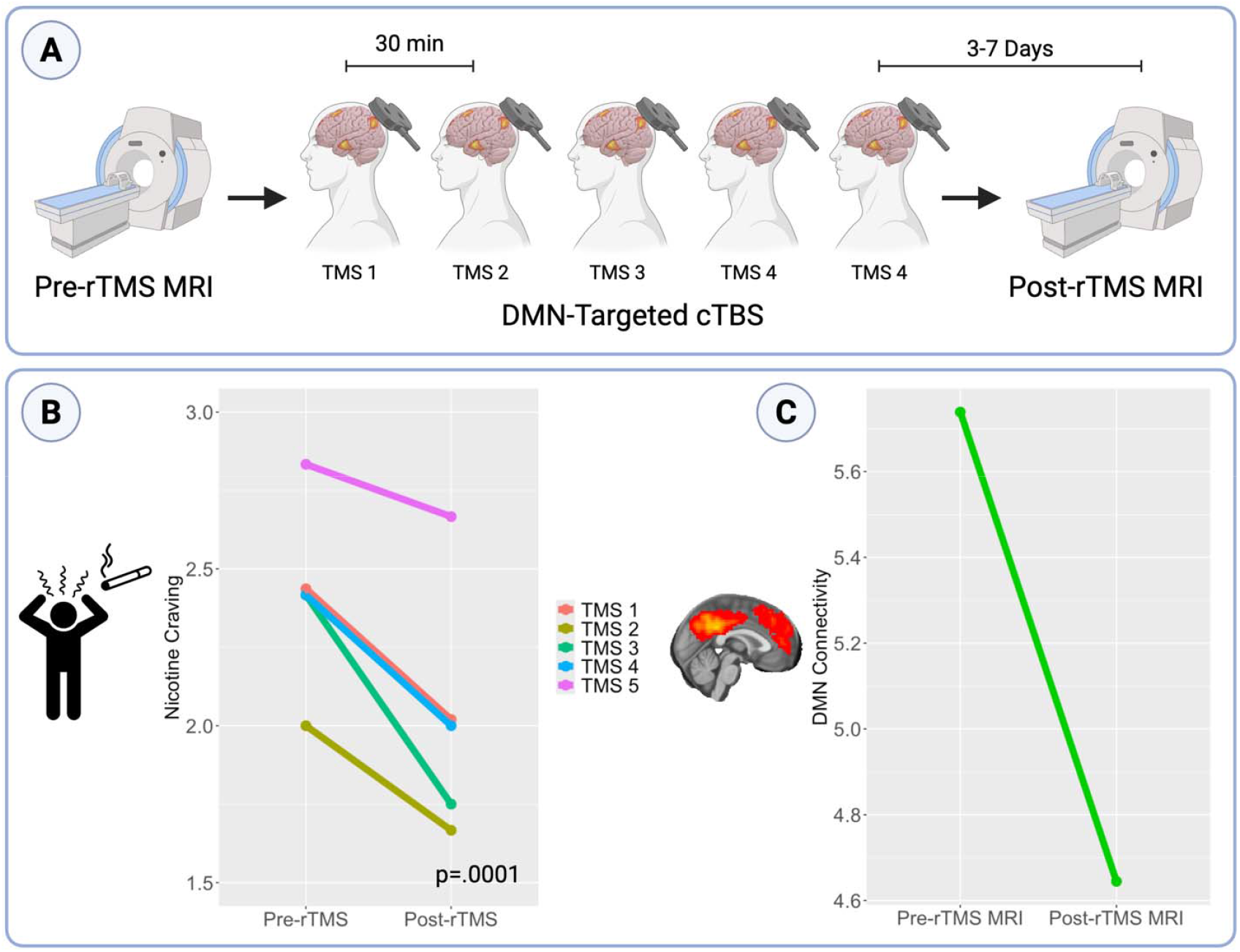
Accelerated Multi-Session DMN-Targeted cTBS Reduces Nicotine Craving and DMN Connectivity. In a study of Accelerated, Multi-Session DMN-targeted cTBS, individuals received five sessions of continuous theta burst stimulation (cTBS) applied to a personalized DMN target separated by 30 minutes with pre- and post-neuroimaging (Figure 4A). Nicotine craving was assessed immediately before and after each single cTBS session using 0-10 visual analog scale. Participants underwent resting-state neuroimaging prior to cTBS and 3-7 days after cTBS. There were significant effects of time (i.e., pre vs. post each rTMS session, p=.0001, Figure 4B) and rTMS session number (p=.0005) on craving, indicating that craving significantly decreased after each session of DMN-targeted cTBS. As evidence of target engagement following DMN-targeted cTBS, we observed a reduction in DMN connectivity that approached significance (p=.052, Figure 4C). Created with BioRender.com.

There were significant effects of time (i.e., post-pre=-0.40, X^2^=14.67, df=1, p=.0001, CI=[-0.61, -0.19], Figure 4B; Supplemental Table 3, Supplemental Figure 7) and rTMS session number (X^2^=19.80, df=4, p=.0005; Supplemental Tables 3-4) on craving, indicating that craving significantly decreased after each session of DMN-targeted cTBS. In the per protocol analysis, craving did not change between the pre- and post-rTMS MRI visits (post-pre=-0.273, t(10)=0.396, p=.70). When we included the participant with a protocol deviation (i.e., whose post-rTMS MRI was 12 days after rTMS instead of ≤ 7 days), our findings were unchanged (post-pre=-0.333, t(11)=0.528, p=.608).

#### Multiple Sessions of DMN-Targeted cTBS Reduces DMN Connectivity

In the per-protocol analysis, we observed a reduction in DMN connectivity that approached significance (t(8)=-2.28, CI=[-2.20, 0.011], p=.052, Figure 4C & Supplemental Figure 8). When we included the participant with a protocol deviation, the effect on DMN connectivity was reduced (t(9)=-0.82, 95% CI -1.97 to 0.32, p=.139). Exploratory analyses of individual DMN regions of interest in supplement.

## Discussion

Despite decades of research, nicotine dependence continues to pose a significant public health threat for individuals with schizophrenia due to the high prevalence of nicotine use and minimally effective smoking cessation treatments in this population. Here we used a large, multisite dataset of individuals with psychotic disorders to establish a novel relationship between DMN connectivity and nicotine use in psychosis-spectrum disorders. In people with psychosis, we observed a relationship where lower DMN connectivity was associated with higher daily cigarette use and more recent tobacco use, suggestive of a dose-response relationship. Our findings suggest a novel mechanism for the high prevalence of nicotine use in this population.

After establishing a relationship between DMN connectivity and nicotine use in psychosis, we targeted this network with single and multiple sessions of rTMS applied to an individualized DMN target. A single session of iTBS increased craving compared to cTBS and sham. Given the hypothesized opposing effects of iTBS and cTBS, we concluded that a single session of cTBS (delivered at 80% AMT) was insufficient to reduce craving, so we next tested multiple sessions of cTBS at a higher stimulation intensity (100% AMT) delivered in an accelerated one-day protocol. Sure enough, with a higher stimulation intensity, we observed *robust* and *reproducible* effects where each session of cTBS reduced craving within- and between-participants. We observed demonstrable evidence of target engagement, as DMN connectivity was reduced following cTBS.

In this study, we have identified a novel mechanism driving the high prevalence of nicotine use in schizophrenia and then used two mechanistic rTMS studies to demonstrate target engagement. Our analysis advances the field in several ways. First, we have now established a relationship between nicotine use and DMN connectivity in psychosis. Although DMN connectivity has been linked to nicotine craving in non-psychosis populations (21-26), previous work targeting the DMN with rTMS in the general population has not proven successful in reducing nicotine craving or withdrawal (27). In contrast, we have now shown that targeting the DMN does affect craving in schizophrenia, providing evidence for an alternative mechanism driving nicotine use in people with psychosis, which may lead to more effective interventions for a population disproportionately at risk for the devastating consequences of their tobacco use. While we did not observe a significant group*tobacco use interaction on DMN connectivity in B-SNIP2, the control population may have been underpowered. Our analyses show promising results of modulating the DMN in psychosis but did not test the effects of modulating DMN connectivity in control populations using nicotine. To test if DMN modulation is more effective for nicotine craving in psychosis populations, future studies should prospectively compare the effects of DMN-targeted rTMS in psychosis and control participants who use nicotine. DMN modulation may offer a more effective target for nicotine use in psychosis.

For decades, the field has sought to understand the high prevalence of nicotine use in schizophrenia by focusing on known overlapping abnormalities in reward circuitry in psychotic disorders and substance use (4). However, this research has not led to more effective smoking cessation interventions in schizophrenia. Four-week continuous abstinence rates in schizophrenia following FDA-approved smoking cessation treatments lag below 20% – half the abstinence rate of the general population (3, 28). Our findings suggest that interventions targeting abnormal DMN connectivity may be more effective for smoking cessation in schizophrenia. Notabaly, our observed effects on craving were brief, suggesting that many more sessions of rTMS will be necessary to produce durable effects on craving and ultimately smoking cessation, consistent with prior studies (29-31). Future studies should test if treatments modulating DMN connectivity improve smoking cessation rates in this population.

To our knowledge, this is the first study of single rTMS sessions in schizophrenia with pre-/post-rTMS neuroimaging. Single-session rTMS studies provide critical information on mechanism but are difficult to execute (27, 32), especially in symptomatic clinical populations, as the effects of a single session of rTMS are fleeting, generally lasting less than 60 minutes (33).

Ours is also the first test of an accelerated rTMS intervention for nicotine use in any population and the first for any substance use disorder in schizophrenia. Only a handful of studies have previously tested accelerated protocols for schizophrenia (34-37) and substance use disorders (38), but there is growing interest in developing accelerated protocols that may be more efficient and shorten time to response (39, 40).

Our analyses have several limitations. Although the B-SNIP2 sample was large (n=596), our two mechanistic rTMS studies were relatively small (n=11 and n=12) and should be replicated in larger samples. However, it should be noted that although the Single-Session rTMS study included neuroimaging data from 10 participants, we employed a within-subjects dense MRI scanning protocol where each participant underwent 7 scans, totaling 270 min/participant, including 140 min of resting-state imaging. Therefore, these participants provided a total of 70 separate scans for analysis, representing the first precision functional mapping study with rTMS in schizophrenia. Importantly, our Accelerated Multi-Session cTBS study lacked a sham control, which will be critical for future studies. Another limitation of our analysis was the assessment of craving using self-reported VAS. While VAS for craving is brief and simple – both critical aspects for these mechanistic rTMS studies – it lacks granularity. This may be why we did not observe an *enduring* effect of our accelerated, multi-session cTBS on craving. Future studies should use more sophisticated assessments of craving, such as in-scanner cue-reactivity tasks, as correlates of target engagement.

In conclusion, these results provide evidence for a novel mechanism of nicotine dependence in psychosis that could lead to the first schizophrenia-specific treatment for nicotine dependence, which would combat nicotine’s devastating effects on morbidity and mortality in this vulnerable population.

## Data Availability

All data produced in the present study are available upon reasonable request to the authors.

## Data Availability

Data is available upon reasonable request.

## Author Contributions

HBW: conceptualization, data curation, formal analysis, funding acquisition, investigation, methodology, project administration, resources, software, supervision, validation, visualization, writing – original draft, and writing – reviewing & editing. SB: investigation, project administration, formal analysis, and writing – reviewing & editing. SV: formal analysis and writing – reviewing & editing. BPR: data curation, formal analysis, and writing – reviewing & editing. GY: project administration, investigation, and writing – reviewing & editing. JC: investigation, project administration, visualization, and writing – reviewing & editing. BC: funding acquisition and writing – reviewing & editing. EG: funding acquisition and writing – reviewing & editing. MK: funding acquisition and writing – reviewing & editing. SM: data curation and writing – reviewing & editing. GP: funding acquisition and writing – reviewing & editing. CT: funding acquisition and writing – reviewing & editing. MH: conceptualization, methodology, data curation, and writing – reviewing & editing. RB: conceptualization, data curation, formal analysis, investigation, methodology, supervision, and writing – reviewing & editing.

## References

1. Brown S, Inskip H, Barraclough B. Causes of the excess mortality of schizophrenia. Br J Psychiatry. 2000;177:212–7.

2. Laursen TM, Nordentoft M, Mortensen PB. Excess early mortality in schizophrenia. Annu Rev Clin Psychol. 2014;10:425–48.

3. Evins AE, West R, Benowitz NL, Russ C, Lawrence D, McRae T, et al. Efficacy and Safety of Pharmacotherapeutic Smoking Cessation Aids in Schizophrenia Spectrum Disorders: Subgroup Analysis of EAGLES. Psychiatr Serv. 2021;72(1):7–15.

4. Ward H, Nemeroff C, Carpenter L, Grzenda A, McDonald W, Rodriguez C, et al. Substance use disorders in schizophrenia: Prevalence, etiology, biomarkers, and treatment. Personalized Medicine in Psychiatry. 2023;39-40(July-August 2023).

5. Ward HB, Brady RO, Halko MA, Lizano P. Noninvasive Brain Stimulation for Nicotine Dependence in Schizophrenia: A Mini Review. Front Psychiatry. 2022;13:824878.

6. Ward HB, Beermann A, Nawaz U, Halko MA, Janes AC, Moran LV, et al. Evidence for Schizophrenia-Specific Pathophysiology of Nicotine Dependence. Front Psychiatry. 2022;13:804055.

7. Brady RO, Jr., Gonsalvez I, Lee I, Ongur D, Seidman LJ, Schmahmann JD, et al. Cerebellar-Prefrontal Network Connectivity and Negative Symptoms in Schizophrenia. Am J Psychiatry. 2019:appiajp201818040429.

8. Huang YZ, Edwards MJ, Rounis E, Bhatia KP, Rothwell JC. Theta burst stimulation of the human motor cortex. Neuron. 2005;45(2):201–6.

9. Killen JD, Fortmann SP. Craving is associated with smoking relapse: findings from three prospective studies. Exp Clin Psychopharmacol. 1997;5(2):137–42.

10. Heatherton TF, Kozlowski LT, Frecker RC, Fagerstrom KO. The Fagerstrom Test for Nicotine Dependence: a revision of the Fagerstrom Tolerance Questionnaire. Br J Addict. 1991;86(9):1119–27.

11. Ashburner J, Friston KJ. Unified segmentation. Neuroimage. 2005;26(3):839–51.

12. Friston KJ, Williams S, Howard R, Frackowiak RS, Turner R. Movement-related effects in fMRI time-series. Magn Reson Med. 1996;35(3):346–55.

13. Behzadi Y, Restom K, Liau J, Liu TT. A component based noise correction method (CompCor) for BOLD and perfusion based fMRI. Neuroimage. 2007;37(1):90–101.

14. Ashburner J. A fast diffeomorphic image registration algorithm. Neuroimage. 2007;38(1):95–113.

15. Foulds J, Veldheer S, Yingst J, Hrabovsky S, Wilson SJ, Nichols TT, et al. Development of a questionnaire for assessing dependence on electronic cigarettes among a large sample of ex-smoking E-cigarette users. Nicotine Tob Res. 2015;17(2):186–92.

16. Cox LS, Tiffany ST, Christen AG. Evaluation of the brief questionnaire of smoking urges (QSU-brief) in laboratory and clinical settings. Nicotine Tob Res. 2001;3(1):7–16.

17. Welsch SK, Smith SS, Wetter DW, Jorenby DE, Fiore MC, Baker TB. Development and validation of the Wisconsin Smoking Withdrawal Scale. Exp Clin Psychopharmacol. 1999;7(4):354–61.

18. Turi Z, Lenz M, Paulus W, Mittner M, Vlachos A. Selecting stimulation intensity in repetitive transcranial magnetic stimulation studies: A systematic review between 1991 and 2020. Eur J Neurosci. 2021;53(10):3404–15.

19. Yeo BT, Krienen FM, Sepulcre J, Sabuncu MR, Lashkari D, Hollinshead M, et al. The organization of the human cerebral cortex estimated by intrinsic functional connectivity. J Neurophysiol. 2011;106(3):1125–65.

20. Raichle ME. The restless brain. Brain Connect. 2011;1(1):3–12.

21. Wang KS, Kaiser RH, Peechatka AL, Frederick BB, Janes AC. Temporal Dynamics of Large-Scale Networks Predict Neural Cue Reactivity and Cue-Induced Craving. Biol Psychiatry Cogn Neurosci Neuroimaging. 2020;5(11):1011–8.

22. Hahn B, Ross TJ, Yang Y, Kim I, Huestis MA, Stein EA. Nicotine enhances visuospatial attention by deactivating areas of the resting brain default network. J Neurosci. 2007;27(13):3477–89.

23. Tanabe J, Nyberg E, Martin LF, Martin J, Cordes D, Kronberg E, et al. Nicotine effects on default mode network during resting state. Psychopharmacology (Berl). 2011;216(2):287–95.

24. Janes AC, Betts J, Jensen JE, Lukas SE. Dorsal anterior cingulate glutamate is associated with engagement of the default mode network during exposure to smoking cues. Drug Alcohol Depend. 2016;167:75–81.

25. Janes AC, Datko M, Roy A, Barton B, Druker S, Neal C, et al. Quitting starts in the brain: a randomized controlled trial of app-based mindfulness shows decreases in neural responses to smoking cues that predict reductions in smoking. Neuropsychopharmacology. 2019;44(9):1631–8.

26. Tabibnia G, Ghahremani DG, Pochon JF, Diaz MP, London ED. Negative affect and craving during abstinence from smoking are both linked to default mode network connectivity. Drug Alcohol Depend. 2023;249:109919.

27. Petersen N, Apostol MR, Jordan T, Ngo TDP, Kearley NW, London ED, et al. Comparing neuromodulation targets to reduce cigarette craving and withdrawal: a randomized clinical trial. Neuropsychopharmacology. 2025.

28. Anthenelli RM, Benowitz NL, West R, St Aubin L, McRae T, Lawrence D, et al. Neuropsychiatric safety and efficacy of varenicline, bupropion, and nicotine patch in smokers with and without psychiatric disorders (EAGLES): a double-blind, randomised, placebo-controlled clinical trial. Lancet. 2016;387(10037):2507–20.

29. Dinur-Klein L, Dannon P, Hadar A, Rosenberg O, Roth Y, Kotler M, et al. Smoking cessation induced by deep repetitive transcranial magnetic stimulation of the prefrontal and insular cortices: a prospective, randomized controlled trial. Biol Psychiatry. 2014;76(9):742–9.

30. Zangen A, Moshe H, Martinez D, Barnea-Ygael N, Vapnik T, Bystritsky A, et al. Repetitive transcranial magnetic stimulation for smoking cessation: a pivotal multicenter double-blind randomized controlled trial. World Psychiatry. 2021;20(3):397–404.

31. Prikryl R, Ustohal L, Kucerova HP, Kasparek T, Jarkovsky J, Hublova V, et al. Repetitive transcranial magnetic stimulation reduces cigarette consumption in schizophrenia patients. Prog Neuropsychopharmacol Biol Psychiatry. 2014;49:30–5.

32. Ward HB. One step at a time: use of single-session rTMS to test novel targets for substance use disorders. Neuropsychopharmacology. 2025.

33. Walther S, Kunz M, Müller M, Zürcher C, Vladimirova I, Bachofner H, et al. Single Session Transcranial Magnetic Stimulation Ameliorates Hand Gesture Deficits in Schizophrenia. Schizophr Bull. 2020;46(2):286–93.

34. Sverak T, Mayerova M, Obdrzalkova M, Ustohal L. Accelerated Repetitive Transcranial Magnetic Stimulation in the Treatment of Negative Symptoms of Schizophrenia: An Open-Label Study. J ECT. 2022;38(2):e24–e5.

35. Kang D, Zhang Y, Wu G, Song C, Peng X, Long Y, et al. The Effect of Accelerated Continuous Theta Burst Stimulation on Weight Loss in Overweight Individuals With Schizophrenia: A Double-Blind, Randomized, Sham-Controlled Clinical Trial. Schizophr Bull. 2024;50(3):589–99.

36. Han Y, Jin F, Lee J, Hou W, Yang X, Zhang Y, et al. Accelerated iTBS with a personalised targeting method to treat negative symptoms of schizophrenia: A randomized controlled trial. Brain Stimul. 2025;18(3):710–9.

37. Jin Y, Tong J, Huang Y, Shi D, Zhu N, Zhu M, et al. Effectiveness of accelerated intermittent theta burst stimulation for social cognition and negative symptoms among individuals with schizophrenia: A randomized controlled trial. Psychiatry Res. 2023;320:115033.

38. Steele VR, Maxwell AM, Ross TJ, Stein EA, Salmeron BJ. Accelerated Intermittent Theta-Burst Stimulation as a Treatment for Cocaine Use Disorder: A Proof-of-Concept Study. Front Neurosci. 2019;13:1147.

39. van Rooij SJH, Arulpragasam AR, McDonald WM, Philip NS. Accelerated TMS - moving quickly into the future of depression treatment. Neuropsychopharmacology. 2024;49(1):128–37.

40. Ward HB, Blyth SH, Kast K. Rewiring recovery: Patient-centered neuromodulation interventions for substance use disorders that meet people where they are. Transcranial Magnetic Stimulation. 2025;4(August 2025).

