## Supplement for "Defining and Engaging a Novel rTMS Target for Nicotine Craving in Psychotic Disorders"

**Supplemental Material**

**Methods**
***B-SNIP2***

*Participants:* Data came from the Bipolar-Schizophrenia Network on Intermediate Phenotypes 2 (B-SNIP2), a multisite, cross-sectional study of individuals with psychotic disorders who underwent clinical characterization, cognitive assessment, neurophysiologic phenotyping, and structural, diffusion tensor, and resting-state functional brain imaging. A blood sample was also collected from each participant for genetic analysis. B-SNIP2 procedures were the same as in B-SNIP1 (1). DSM-IV-TR diagnoses were made at consensus meetings using all available information, including findings from the Structured Clinical Interview for DSM-IV Axis I Disorders (SCID-IV) (2) administered by trained clinical raters who held monthly inter-site reliability conference calls (1). All individuals with psychosis were on stable medication with no major changes in the past 30 days. Healthy control participants had no history of a psychotic disorder or recurrent mood disorder, and no known close relatives with these disorders. Exclusion criteria included history of head injury with loss of consciousness > 10 min; pregnancy; positive urine toxicology for drugs of abuse on the day of testing; diagnosis of substance abuse in the past 30 days or substance dependence in the past 3 months; history of systemic medical or neurological disorder affecting mood or cognition; or intellectual disability. The final sample (Table 1) included 276 healthy individuals and 320 individuals with a psychotic disorder (schizophrenia = 129, schizoaffective disorder = 124, bipolar disorder with psychotic features = 67).

*MRI Acquisition and Data Processing:* Subjects underwent a 3D T1-weighted structural scan and a single resting-state fMRI (rs-fMRI) 8-minute scan on a 3T scanner. Participants were instructed to keep their eyes open, focus on a crosshair displayed on a monitor, and to remain still during the entire scan. Head motion was minimized with a custom-built head-coil cushion. Scanning protocols across sites are listed in Supplemental Table 1. Preprocessing was performed using the advanced DPARSF module within DPABI V6.0_210501. For each site separately, all volume slices were corrected for different signal acquisition times. Then, the time series of images for each subject were realigned. Individual structural images (T1-weighted) were co-registered to the mean functional image after realignment. The transformed structural images were then segmented into GM, WM and CSF (3). To remove the nuisance signals, the Friston 24-parameter model (4) was utilized to regress out head motion effects from the realigned data. To better control for physiological noise, a combined WM/CSF mask was generated from which the top 5 principal components were extracted and signal adjusted using the CompCor method (5). In addition, linear trends were adjusted since the BOLD signal exhibits low-frequency drifts. The DARTEL tool (6) was used to transform the functional data from individual native space to MNI space. Following this, temporal filtering (0.01–0.08 Hz) was performed.

*Neuroimaging Analysis - Calculation of Whole-Network Connectivity Values:* We calculated individual values of average DMN functional connectivity by placing 6mm spheres at coordinates corresponding to seven standard nodes of the DMN (posterior cingulate/precuneus, medial prefrontal, left lateral parietal, right lateral parietal, left inferior temporal, right inferior temporal, medial dorsal thalamus) (see (7) for coordinates). The time courses of the blood-oxygen-level-dependent (BOLD) signal from the regions of interest (ROIs) were correlated with each other and z-transformed to generate a 7 x 7 ROI to ROI connectivity matrix. A mean connectivity from this matrix was generated for each participant by averaging connectivity values for the entire DMN.

***Single-Session DMN-Targeted rTMS in Schizophrenia***

*Participants:* Diagnosis of schizophrenia or schizoaffective disorder was confirmed by DSM-V SCID interview (8) and clinical information obtained from outpatient psychiatric providers. For one month prior to enrollment, individuals received outpatient care, with no hospitalizations or changes to their psychiatric medication regimens. Individuals were excluded if they had DSM-V intellectual disability, substance use disorder (other than nicotine) in the past 3 months, a progressive or genetic neurologic disorder, history of significant head trauma, history of seizures or neurosurgical procedures, implanted devices, gross organic pathology on neuroimaging, contraindications to MRI or rTMS, or current pregnancy. All participants provided written informed consent in accordance with the Beth Israel Deaconess Medical Center Institutional Review Board.

*MRI Data Processing:* Anatomical images were segmented into gray matter, white matter and cerebrospinal fluid (CSF) with the Computational Anatomy Toolbox 12 (CAT12, version 12.5; [http://www.neuro.uni-jena.de/cat/](https://nam12.safelinks.protection.outlook.com/?url=http%3A%2F%2Fwww.neuro.uni-jena.de%2Fcat%2F&data=05%7C01%7Cheather.b.ward%40vumc.org%7C53146c81d5534edf7b5208db2189eebc%7Cef57503014244ed8b83c12c533d879ab%7C0%7C0%7C638140648518151829%7CUnknown%7CTWFpbGZsb3d8eyJWIjoiMC4wLjAwMDAiLCJQIjoiV2luMzIiLCJBTiI6Ik1haWwiLCJXVCI6Mn0%3D%7C3000%7C%7C%7C&sdata=Fsce8FRawoYi0jL0FjzH%2BUsdG8P5cAj4Wzyuk79pHuo%3D&reserved=0)). Resting-state scans were preprocessed in SPM12 and were (1) realigned to a mean scan, (2) coregistered with the native space structural scan, then (3) underwent resting-state denoising procedures: bandpass filter (0.01–0.1 Hz), regression of CSF and white matter signal, regression of 12 motion parameters (6 translation and rotation parameters and their first derivative). All resting-state scans went through a quality assurance procedure that included calculating framewise displacement (FD) and temporal signal to noise ratio (tSNR). Scans with a mean FD > 0.5 or a tSNR lower than the 5th percentile of the distribution of the entire sample were excluded from further analysis.

*rTMS Protocol:* Individuals participated in a randomized, controlled, crossover study of three single sessions of theta-burst stimulation (Figure 1B and Supplemental Figure 1) applied to an individualized DMN target (see *Individualized DMN Target* below and Supplemental Figure 3) with neuroimaging collected immediately before and after each rTMS session. Baseline anatomical and functional MRIs were used in a Brainsight frameless stereotaxic system (Rogue Research, Montreal, Canada) to target an individualized left lateral parietal DMN region. Frameless stereotaxy was used during all stimulation sessions to monitor the position of the coil throughout rTMS administration. Participants were seated in a chair with their head tilted downward. Individuals received three single sessions of rTMS on three separate days, separated by at least 2 days (48 hours) to avoid any carryover effect (median 6.5 days, mean 13.8 days (SD 22.2), range 2-96 days). Active motor threshold (AMT) was determined prior to the first rTMS session. Participants received one session of iTBS (600 pulses, 100% AMT), one session of cTBS (600 pulses, 80% AMT), and one session of sham (coil flipped 180 degrees using 100% AMT iTBS protocol, 600 pulses). rTMS session order was randomly assigned. rTMS was applied using a MagPro X100 stimulator and an active figure-of-8 coil (Cool B65, MagVenture, Denmark) held tangentially to the scalp with the handle at 45 degrees. rTMS was applied in the standard theta-burst pattern described by Huang et al. (3 pulses at 50-Hz repeated at a rate of 5-Hz) (9). Participants were blinded to rTMS stimulation type. To assess the integrity of the blind, participants were asked after each rTMS session what type of rTMS they thought they received.

*rTMS Protocol Motor Threshold Determination:* Participants had motor threshold determination at their first rTMS visit. Single pulse and repetitive stimulation was performed with a MagPro stimulator (MagVenture) equipped with a biphasic figure-of-eight coil. To obtain an active motor threshold, single pulses were used in the following manner: Electromyographic activity (EMG) was recorded using surface electrodes attached to the skin to measure motor evoked potentials (MEP) during the motor threshold assessment. The TMS coil was placed on the scalp. Single TMS pulses were applied over the hand area of the left motor cortex and individually localized for each participant based on the optimal position for eliciting a motor evoked potential. Neuronavigation (Brainsight, Rogue Research, Inc.) was used to record the motor ‘hot-spot.’ Resting and active motor threshold (RMT; AMT) were obtained by following recommendations from the International Federation of Clinical Neurophysiology.

*Neuroimaging Analysis - Calculation of Whole-Network Connectivity Values:* We calculated the average connectivity of the entire DMN as previously (10). We generated individual values of average DMN functional connectivity by placing 6mm spheres at coordinates corresponding to nine standard nodes of the DMN (posterior cingulate/precuneus, medial prefrontal, left lateral parietal, right lateral parietal, left inferior temporal, right inferior temporal, medial dorsal thalamus, right posterior cerebellum, left posterior cerebellum) (see (7) for coordinates). The time courses of the blood-oxygen-level-dependent (BOLD) signal from the regions of interest (ROIs) were correlated with each other and z-transformed to generate a 9 x 9 ROI to ROI connectivity matrix. A mean connectivity from this matrix was generated for each participant by averaging connectivity values for the entire DMN.

***Accelerated Multi-Session DMN-Targeted rTMS in Schizophrenia***

*Participants:* Diagnosis of schizophrenia or schizoaffective disorder was confirmed by DSM-V SCID interview (8) and clinical information obtained from outpatient psychiatric providers. For one month prior to enrollment, individuals received outpatient care, with no hospitalizations or changes to their psychiatric medication regimens. Individuals were excluded if they had DSM-V intellectual disability, substance use disorder (other than nicotine) in the past 3 months, a progressive or genetic neurologic disorder, history of significant head trauma, history of seizures or neurosurgical procedures, implanted devices, gross organic pathology on neuroimaging, contraindications to MRI or rTMS, or current pregnancy. All participants provided written informed consent in accordance with the Vanderbilt University Medical Center Institutional Review Board.

*MRI Acquisition and Data Processing:* Anatomical images were segmented into gray matter, white matter and cerebrospinal fluid (CSF) with the Computational Anatomy Toolbox 12 (CAT12, version 12.5; [http://www.neuro.uni-jena.de/cat/](https://nam12.safelinks.protection.outlook.com/?url=http%3A%2F%2Fwww.neuro.uni-jena.de%2Fcat%2F&data=05%7C01%7Cheather.b.ward%40vumc.org%7C53146c81d5534edf7b5208db2189eebc%7Cef57503014244ed8b83c12c533d879ab%7C0%7C0%7C638140648518151829%7CUnknown%7CTWFpbGZsb3d8eyJWIjoiMC4wLjAwMDAiLCJQIjoiV2luMzIiLCJBTiI6Ik1haWwiLCJXVCI6Mn0%3D%7C3000%7C%7C%7C&sdata=Fsce8FRawoYi0jL0FjzH%2BUsdG8P5cAj4Wzyuk79pHuo%3D&reserved=0)). Resting-state scans were preprocessed in SPM12 and were (1) realigned to a mean scan, (2) coregistered with the native space structural scan, then (3) underwent resting-state denoising procedures: bandpass filter (0.01–0.1 Hz), regression of CSF and white matter signal, regression of 12 motion parameters (6 translation and rotation parameters and their first derivative). All resting-state scans went through a quality assurance procedure that included calculating framewise displacement (FD) and temporal signal to noise ratio (tSNR). Scans with a mean FD > 0.2 were excluded from further analysis.

*rTMS Protocol Motor Threshold Determination:* Participants had motor threshold determination at their first rTMS visit. Single pulse and repetitive stimulation was performed with a MagPro stimulator (MagVenture) equipped with a biphasic figure-of-eight coil. To obtain an active motor threshold, single pulses were used in the following manner: Electromyographic activity (EMG) was recorded using surface electrodes attached to the skin to measure motor evoked potentials (MEP) during the motor threshold assessment. The TMS coil was placed on the scalp. Single TMS pulses were applied over the hand area of the left motor cortex and individually localized for each participant based on the optimal position for eliciting a motor evoked potential. Neuronavigation (Brainsight, Rogue Research, Inc.) was used to record the motor ‘hot-spot.’ Resting and active motor threshold (RMT; AMT) were obtained by following recommendations from the International Federation of Clinical Neurophysiology.

*Neuroimaging Analysis - Calculation of Whole-Network Connectivity Values:* We calculated the average connectivity of the entire DMN as previously (10). We generated individual values of average DMN functional connectivity by placing 6mm spheres at coordinates corresponding to nine standard nodes of the DMN (posterior cingulate/precuneus, medial prefrontal, left lateral parietal, right lateral parietal, left inferior temporal, right inferior temporal, medial dorsal thalamus, right posterior cerebellum, left posterior cerebellum) (see (7) for coordinates). The time courses of the blood-oxygen-level-dependent (BOLD) signal from the regions of interest (ROIs) were correlated with each other and z-transformed to generate a 9 x 9 ROI to ROI connectivity matrix. A mean connectivity from this matrix was generated for each participant by averaging connectivity values for the entire DMN.

***Individualized DMN Targeting****:* The DMN target was identified using the same methods for the Single-Session rTMS and Accelerated Multi-Session rTMS studies. For our DMN target, we selected the left lateral parietal DMN, as it is a DMN region that is readily identifiable across all individuals and has been successfully used to modulate DMN connectivity with rTMS (11). To identify an individualized DMN map for rTMS targeting, a standard DMN template (12) was warped into native space and applied to the participant’s baseline or pre-rTMS scan. In each participant, the resultant connectivity maps yielded a correlation cluster in the left posterior inferior parietal lobule (IPL, Supplemental Figure 3). A target was then placed in the averaged center of the left posterior IPL correlation cluster (formed from the overlay of the left posterior IPL clusters derived from the connectivity maps) on the cortical surface using Brainsight neuronavigation software (Rogue Research, Inc.) An individualized rTMS target was selected in the left parietal region of the DMN and used as the rTMS target for all rTMS sessions.

**Results**
***B-SNIP2***

*Individuals with Psychosis are More Likely to Use Tobacco and are More Nicotine-Dependent*

In the B-SNIP2 sample, individuals with psychosis were more likely to have ever used tobacco (X^2^ = 81.816, df = 1, p<2.2e-16, Supplemental Figure 4A) than controls. Among current tobacco users, the psychosis group was more nicotine-dependent than controls (FTND 2.0 vs. 3.312, p=.0004). Individuals in the psychosis group were more likely than controls to have used tobacco in the last 30 days (65.8% psychosis vs. 30.2% controls, X^2^ = 24.495, df = 1, p=7.451e-7, Supplemental Figure 4B) and in the last 24 hours (35.8% psychosis vs. 5.5% controls, X^2^ = 78.161, df = 1, p<2.2e-16, Supplemental Figure 4C).

***Single-Session DMN-Targeted rTMS***

*Single-Session DMN-Targeted rTMS is Safe and Well-Tolerated in Schizophrenia*

rTMS was safe and well-tolerated. All 10 participants who were randomized completed all three rTMS/fMRI visits. No serious adverse events were observed. Participants endorsed the following side effects: headache or neck pain (n=4) and trouble concentrating (n=5). One participant experienced hypomania that responded to medication adjustment. Four participants reported lower anxiety, and one participant reported improved mood.

After each rTMS single session, participants were asked what type of rTMS they thought they received: cTBS, iTBS, or sham. Of 30 total rTMS sessions, in 14 of them (46.67%) the participant correctly guessed the type of rTMS they received. Of 10 sham sessions, in 4 of them (40%), participants correctly guessed that they were receiving sham.

*Craving is Associated with DMN Connectivity*

Across all participants and sessions, pre-rTMS craving (VAS) was associated with average connectivity of the entire DMN (r=0.49, p=.0097) (Figure 3B). Withdrawal was significantly related to pre-rTMS craving assessed via VAS (r=0.43, p=.018) and Tiffany QSU (r=0.42, p=.0062). Average within-DMN connectivity change did not correlate with craving change (r=-0.17, p=.42).

*State-Dependent Effects: Pre-rTMS Craving Does Not Predict Neural Response*

Given the known state-dependent effects of rTMS, we investigated if pre-rTMS craving assessed via VAS predicted connectivity change. After controlling for craving change, pre-rTMS craving did not significantly predict connectivity change (left-right parietal connectivity change: F=1.2619, df1=1, df2=18.620, p=.28; left parietal-dACC connectivity change: F=0.000, df1=1, df2=19.157, p=.9955).

***Accelerated Multi-Session DMN-Targeted rTMS***

*Accelerated Multi-Session DMN-Targeted rTMS is Safe and Well-Tolerated in Schizophrenia*

rTMS was safe and well-tolerated. All 12 participants received all 5 sessions of cTBS and underwent pre-/post-rTMS neuroimaging. No serious adverse events were observed. One participant reported mild anxiety and low mood after the first rTMS session and slight irritability the day after rTMS.

*Multiple Sessions of DMN-Targeted cTBS Reduces DMN Connectivity*

As an exploratory analysis in individuals who completed MRI scans per-protocol, we tested if the connectivity between any individual DMN regions of interest were affected by cTBS. We observed significant reductions in connectivity between the left lateral parietal rTMS target and the 1) dorsomedial prefrontal cortex (t(8)=-2.78, p=.02) and 2) right posterior cerebellum (t(8)=-2.47, p=.04).

**Supplemental Tables & Figures:**

**Supplemental Table 1. BSNIP2 resting-state fMRI Acquisition Parameters**

| **Site** | **TR (ms)** | **TE (ms)** | **FA (degree)** | **Slices (N)** | **slice order** | **Acquisition matrix (mm)** | **Voxel Size (mm)** | **Vendor** |
| --- | --- | --- | --- | --- | --- | --- | --- | --- |
| **Georgia** | 2000ms | 30ms | 60 | 30 | Interleaved ascending | 64x64 | 3.4x3.4x5 | GE HDx |
| **Hartford** | 2000ms | 30ms | 60 | 30 | Sequential ascending | 64x64 | 3.4x3.4x5 | Siemens Skyra |
| **Boston** | 2000ms | 30ms | 60 | 30 | Sequential ascending | 64x64 | 3.4x3.4x4 | GE HDxt |
| **Dallas** | 2000ms | 30ms | 60 | 30 | Sequential ascending | 64x64 | 3.4x3.4x4 | Philips Achieva |
| **Chicago** | 2000ms | 30ms | 60 | 30 | Sequential ascending | 64x64 | 3.4x3.4x4 | Philips dStream Achieva |

**Supplemental Table 2. B-SNIP2 Pairwise Comparisons of DMN Connectivity by Tobacco Use History**

| **Comparison** | **Estimate** | **SE** | **df** | **t** | **p** |
| --- | --- | --- | --- | --- | --- |
| ***Entire Sample*** | | | | | |
| Current - Former | -0.04022 | 0.0171 | 583 | -2.351 | 0.0191 |
| Current - Never | -0.03151 | 0.0136 | 583 | -2.310 | 0.0212 |
| Former - Never | 0.00871 | 0.0154 | 583 | 0.565 | 0.5720 |
| ***Psychosis*** | | | | | |
| Current - Former | -0.04606 | 0.0213 | 308 | -2.162 | 0.0314 |
| Current - Never | -0.04371 | 0.0182 | 308 | -2.403 | 0.0169 |
| Former - Never | 0.00234 | 0.0222 | 308 | 0.106 | 0.9158 |
| ***Controls*** | | | | | |
| Current - Former | -0.0229 | 0.0363 | 266 | -0.630 | 0.5291 |
| Current - Never | -0.0116 | 0.0323 | 266 | -0.359 | 0.7201 |
| Former - Never | 0.0113 | 0.0224 | 266 | 0.503 | 0.6151 |

**Supplemental Table 3. Accelerated, Multi-Session DMN-Targeted cTBS Craving Effects by TMS Session**

| **Session** | **Estimate** | **SE** | **df** | **t** | **p** |
| --- | --- | --- | --- | --- | --- |
| TMS 1 Post - Pre | -0.417 | 0.452 | 99 | 0.923 | 0.3584 |
| TMS 2 Post - Pre | -0.333 | 0.188 | 99 | 1.773 | 0.0793 |
| TMS 3 Post - Pre | -0.667 | 0.284 | 99 | 2.345 | 0.0210 |
| TMS 4 Post - Pre | -0.417 | 0.149 | 99 | 2.803 | 0.0061 |
| TMS 5 Post - Pre | -0.167 | 0.167 | 99 | 1.000 | 0.3197 |

**Supplemental Table 4. Accelerated, Multi-Session DMN-targeted cTBS Effects on Craving Post-Hoc Testing**

| **Visit** | **Estimate** | **SE** | **df** | **Lower CL** | **Upper CL** |
| --- | --- | --- | --- | --- | --- |
| TMS 1 | 2.21 | 0.562 | 14.8 | 1.009 | 3.41 |
| TMS 2 | 1.83 | 0.437 | 14.8 | 0.902 | 2.76 |
| TMS 3 | 2.08 | 0.564 | 14.8 | 0.881 | 3.29 |
| TMS 4 | 2.21 | 0.608 | 14.8 | 0.912 | 3.50 |
| TMS 5 | 2.75 | 0.641 | 14.8 | 1.382 | 4.12 |

**Supplemental Table 5. Post-Hoc Testing of Accelerated, Multi-Session DMN-Targeted cTBS Effects on Craving – Between-Session Contrasts**

| **Contrast** | **Estimate** | **SE** | **df** | **t** | **p** |
| --- | --- | --- | --- | --- | --- |
| TMS 1-TMS 2 | 0.375 | 0.354 | 103 | 1.059 | 0.2922 |
| TMS 1-TMS 3 | 0.125 | 0.481 | 103 | 0.260 | 0.7956 |
| TMS 1-TMS 4 | 0.000 | 0.590 | 103 | 0.000 | 1.0000 |
| TMS 1-TMS 5 | -0.542 | 0.687 | 103 | -0.789 | 0.4320 |
| TMS 2-TMS 3 | -0.250 | 0.351 | 103 | -0.713 | 0.4778 |
| TMS 2-TMS 4 | -0.375 | 0.418 | 103 | -0.897 | 0.3717 |
| TMS 2-TMS 5 | -0.917 | 0.456 | 103 | -2.011 | 0.0469* |
| TMS 3-TMS 4 | -0.125 | 0.186 | 103 | -0.672 | 0.5029 |
| TMS 3-TMS 5 | -0.667 | 0.304 | 103 | -2.196 | 0.0303* |
| TMS 4-TMS 5 | -0.542 | 0.250 | 103 | -2.169 | 0.0324* |

*p<.05

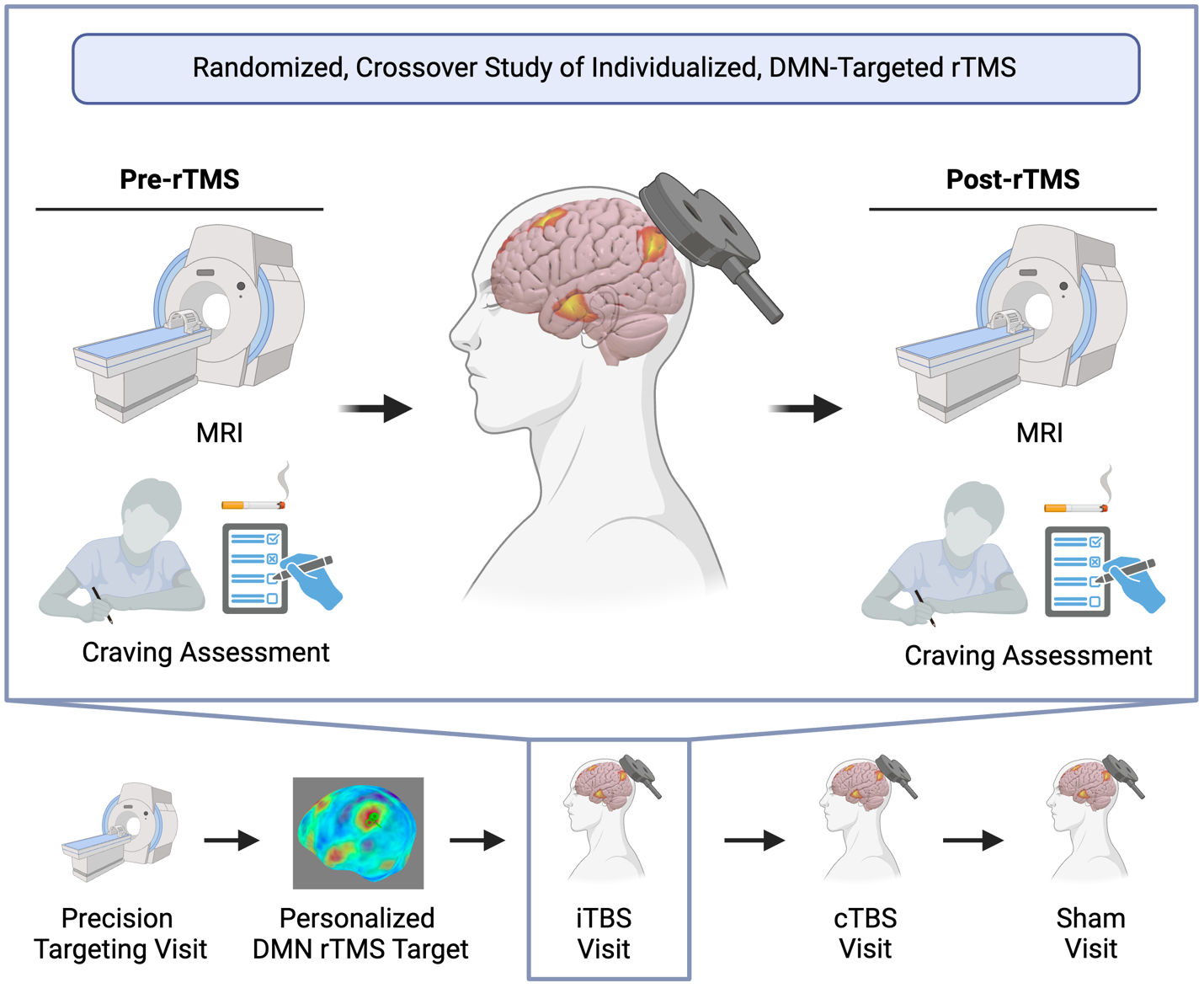

**Supplemental Figure 1. Single-Session DMN-Targeted rTMS Study Design.** We performed a randomized, crossover study of rTMS targeted to an individual-specific map of the left lateral parietal node of the Default Mode Network (DMN). Individuals first completed a Precision Targeting Visit to enable personalized rTMS target selection. Ten individuals with schizophrenia who used nicotine underwent three single sessions of rTMS (intermittent theta burst stimulation, iTBS; continuous theta burst stimulation, cTBS; and sham) in a randomized, crossover design. At each rTMS visit, participants performed behavioral assessments and underwent resting-state fMRI immediately before and after receiving rTMS. Created with BioRender.com.

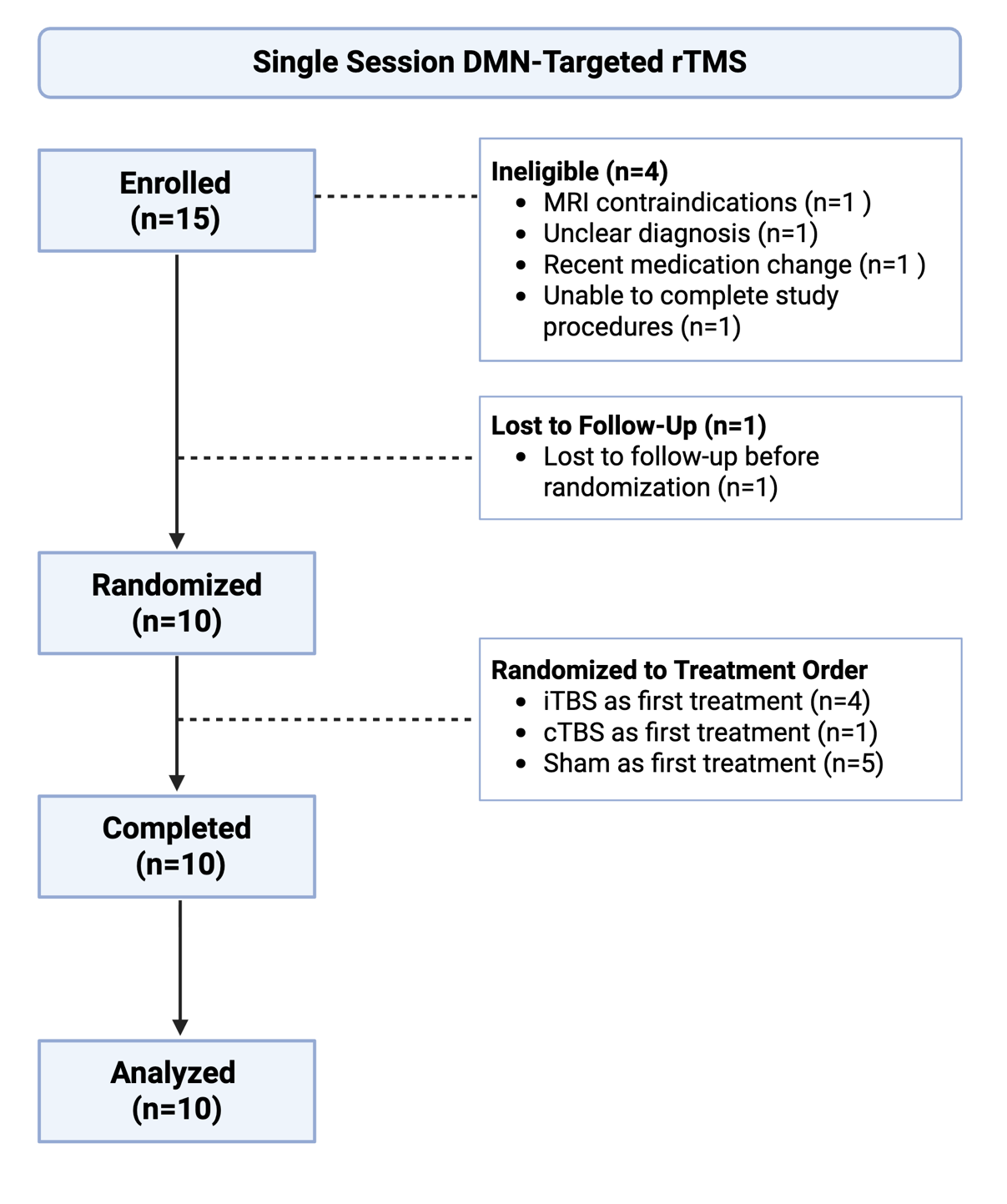

**Supplemental Figure 2. Single-Session DMN-Targeted rTMS CONSORT Diagram.** Fifteen participants with schizophrenia or schizoaffective disorder aged 18-65 who use nicotine were enrolled in this randomized, sham-controlled crossover study. Ten participants completed the study and provided data for analysis.

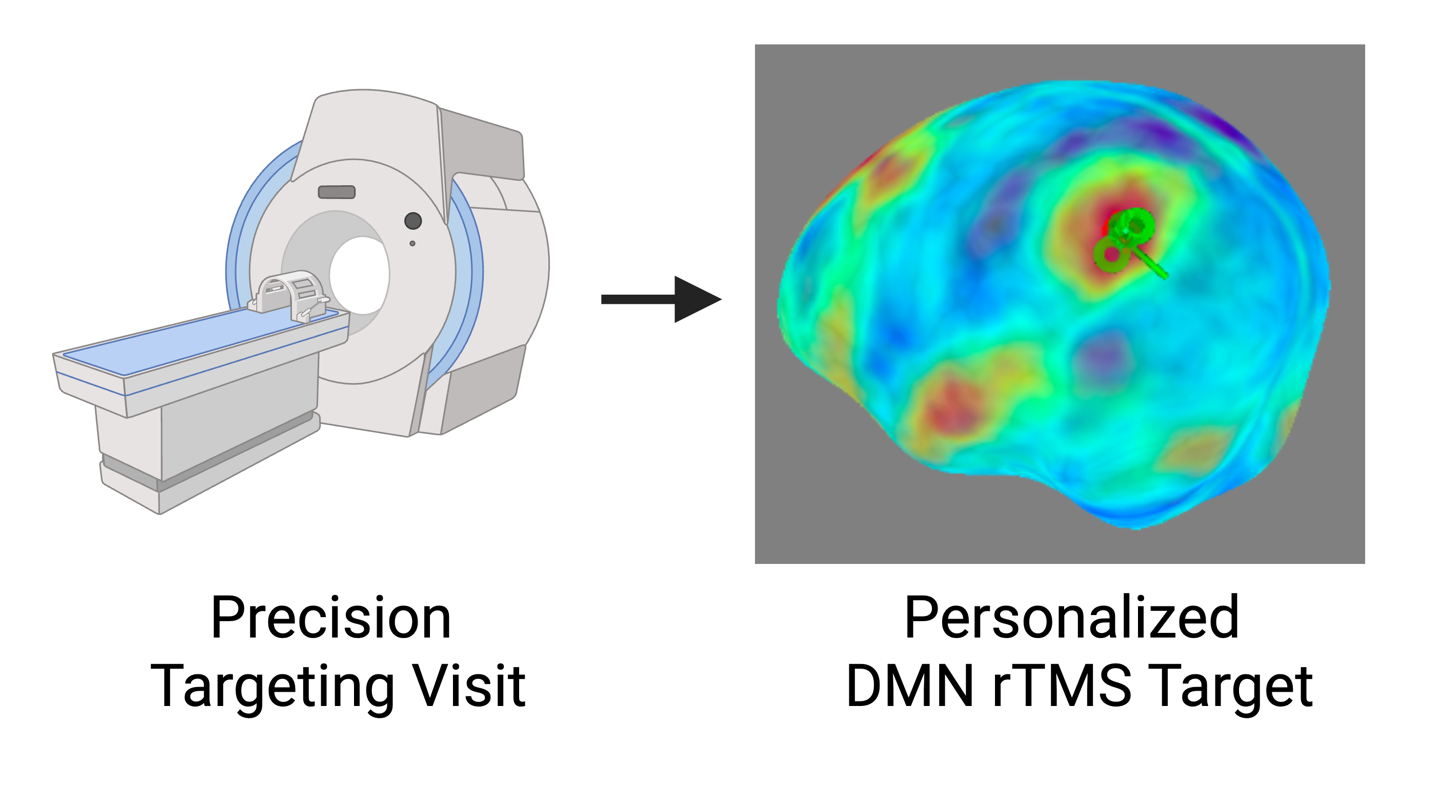

**Supplemental Figure 3. Individualized Default Mode Network rTMS Target***:* The Default Mode Network (DMN) target was identified using the same methods for the Single-Session rTMS and Accelerated Multi-Session rTMS studies. For our DMN target, we selected the left lateral parietal DMN, as it is a DMN region that is readily identifiable across all individuals and has been successfully used to modulate DMN connectivity with rTMS (11). To identify an individualized DMN map for rTMS targeting, a standard DMN template (12) was warped into native space and applied to the participant’s baseline or pre-rTMS scan. The participant scan and DMN mask were then warped back into standard MNI space. In each participant, the resultant connectivity maps yielded a correlation cluster in the left posterior inferior parietal lobule. A target was then placed in the averaged center of the left posterior inferior parietal lobule correlation cluster (formed from the overlay of the left posterior inferior parietal lobule clusters derived from the connectivity maps) on the cortical surface using Brainsight neuronavigation software (Rogue Research, Inc.) An individualized rTMS target was selected in the left parietal region of the DMN and used as the rTMS target for all rTMS sessions. Created with BioRender.com.

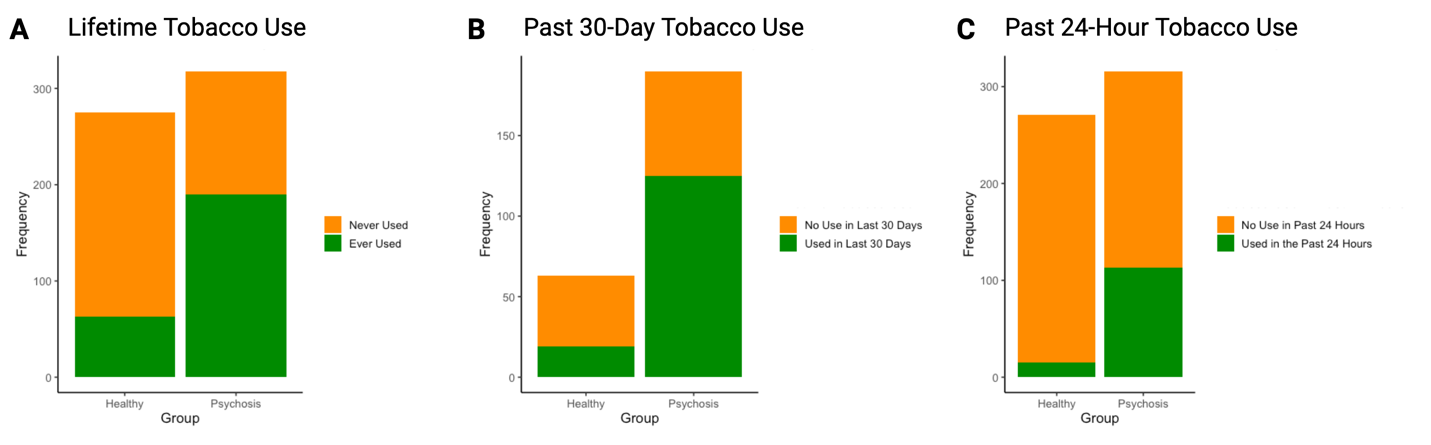

**Supplemental Figure 4. Individuals with Psychosis are More Likely to be Current Tobacco Users than Controls.** Individuals with psychosis were more likely to have ever used tobacco (X^2^ = 81.816, df = 1, p<2.2e-16, Supplemental Figure 4A) than controls. Individuals in the psychosis group were more likely than controls to have used tobacco in the last 30 days (X2 = 24.495, df = 1, p=7.451e-7, Supplemental Figure 4B) and in the last 24 hours (X^2^ = 78.161, df = 1, p<2.2e-16, Supplemental Figure 4C).

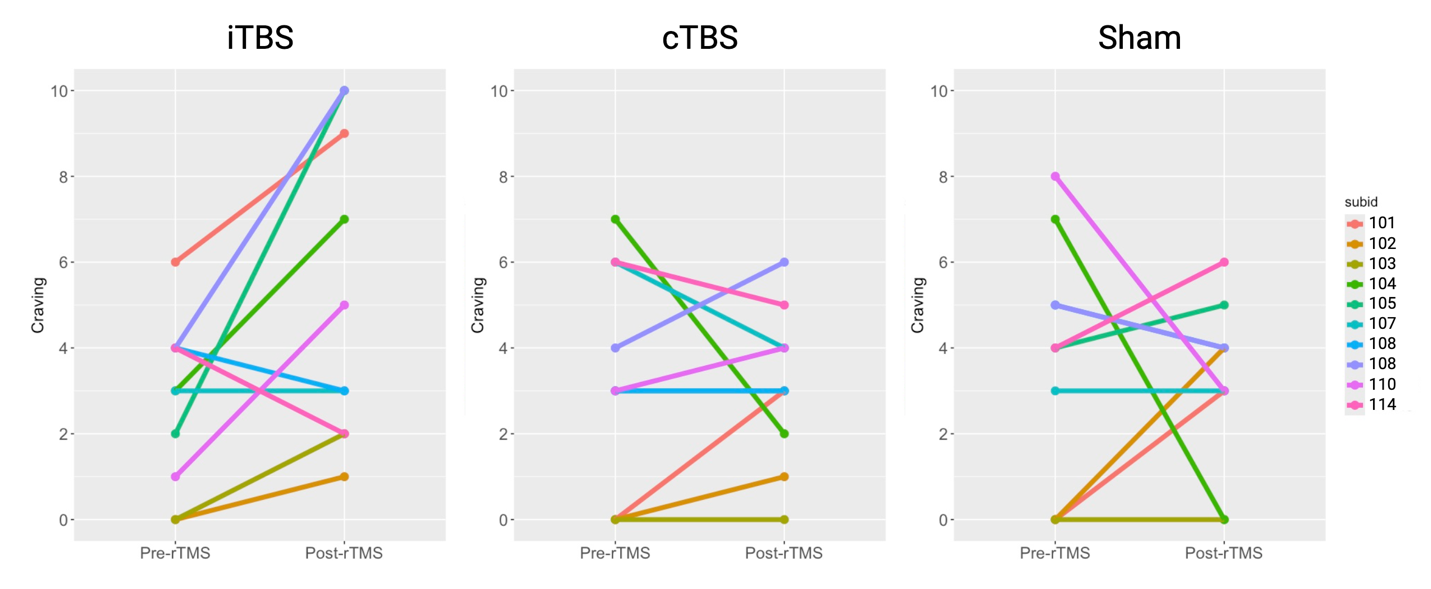

**Supplemental Figure 5. Single-Session DMN Targeted rTMS Subject-Specific Plots of Nicotine Craving Change by rTMS Intervention.** Ten individuals with schizophrenia and daily nicotine use underwent three single sessions of DMN-targeted rTMS (iTBS, cTBS, and sham) with nicotine craving assessment (visual analog scale) immediately before (Pre-rTMS) and after (Post-rTMS) each rTMS session. Shown here are the changes in nicotine craving for each subject shown by rTMS session.

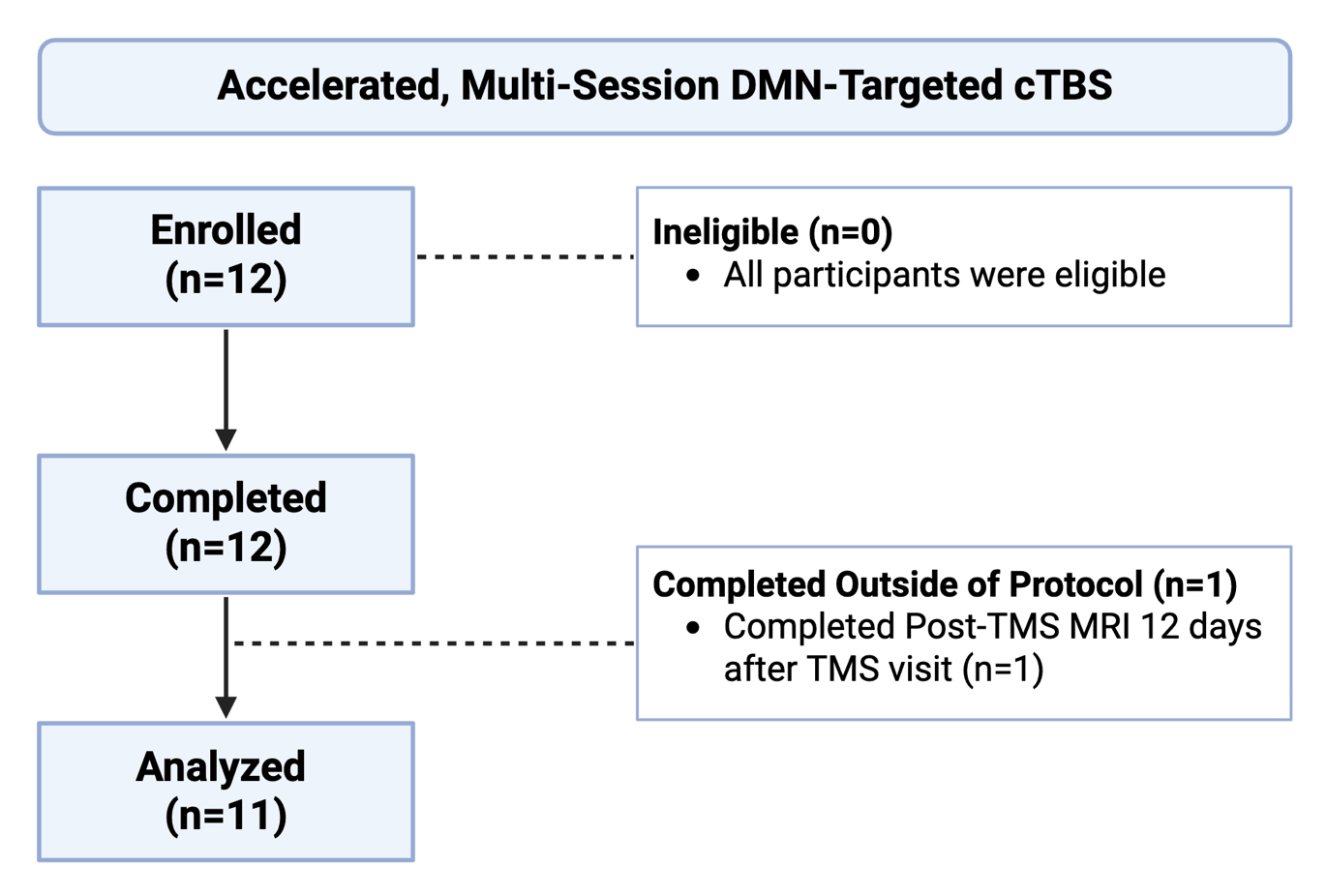

**Supplemental Figure 6. Accelerated, Multi-Session DMN-Targeted cTBS CONSORT Diagram.** Twelve participants with schizophrenia or schizoaffective disorder aged 18-65 who use nicotine were enrolled in this open-label study. Twelve participants completed the study and provided data for analysis. One participant completed the Post-TMS MRI outside of the protocol.

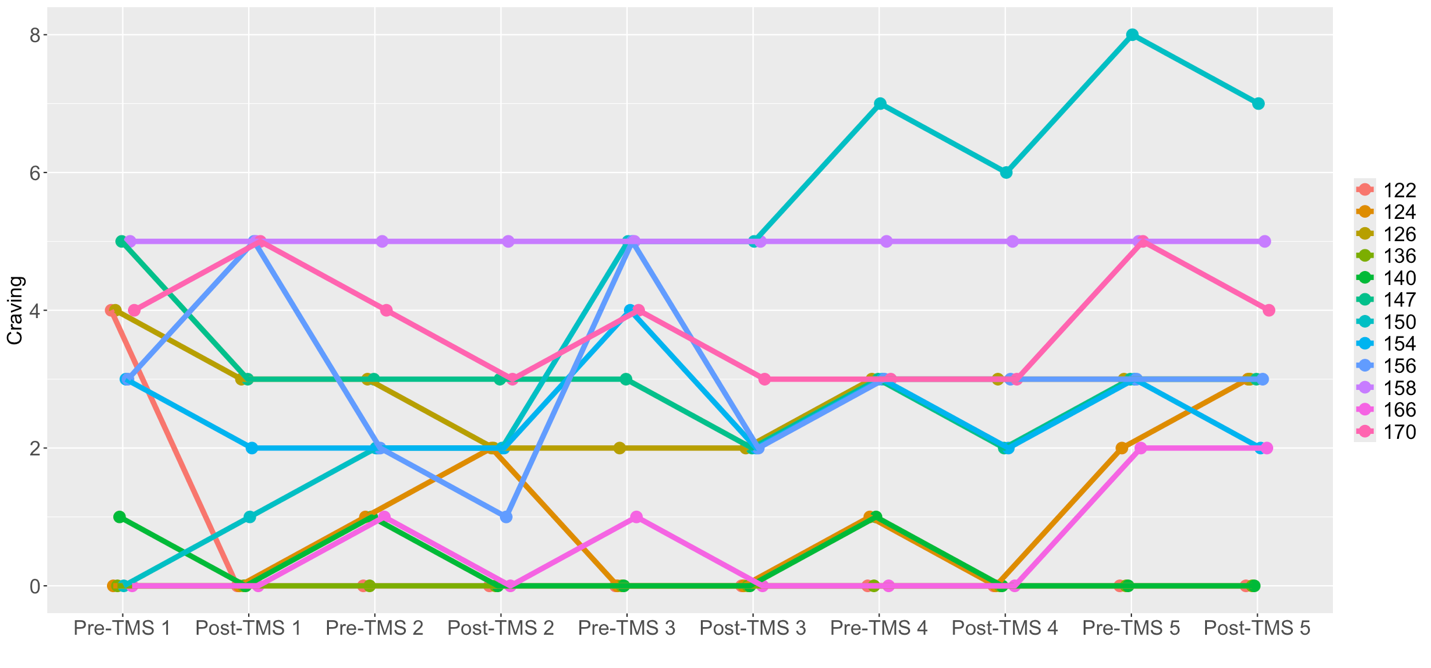

**Supplemental Figure 7. Accelerated, Multi-Session DMN-Targeted cTBS Subject-Specific Plots of Craving Change.** Twelve individuals with schizophrenia and nicotine use underwent 5 sessions of DMN-targeted continuous theta burst stimulation (cTBS) in a one-day accelerated protocol with pre- and post-rTMS neuroimaging. There were significant effects of time (i.e., post-pre each TMS session, p=.0001, Figure 4B, Supplemental Table 3) and TMS session number (p=.0005; Supplemental Tables 3-4) on craving, indicating that craving significantly decreased after each session of DMN-targeted cTBS.

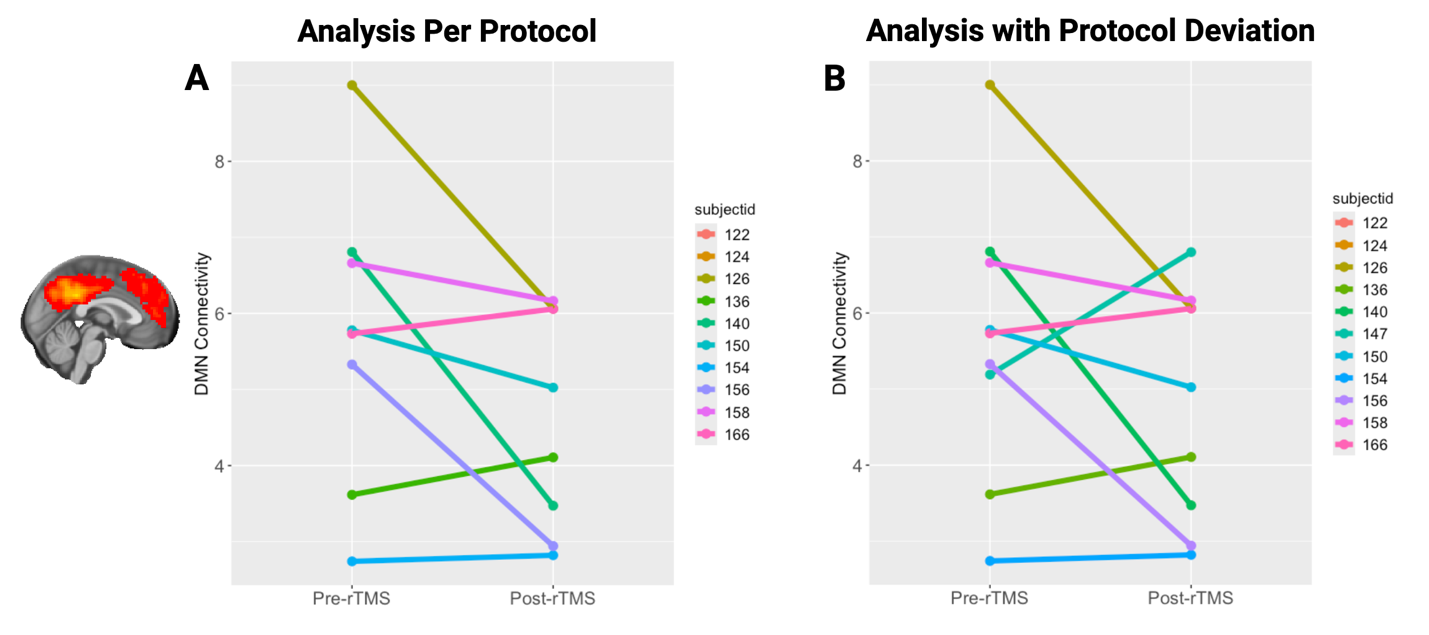

**Supplemental Figure 8. Accelerated, Multi-Session DMN Targeted cTBS Subject-Specific Plots of DMN Connectivity Change.** Twelve individuals with schizophrenia and nicotine use underwent 5 sessions of DMN-targeted continuous theta burst stimulation (cTBS) in a one-day accelerated protocol with pre- and post-rTMS neuroimaging. After quality control, there were 10 participants with MRI data for analysis (20 scans). In the per-protocol analysis, we observed a reduction in DMN connectivity that approached significance (t(8)=-2.28, 95% CI=[-2.20, 0.011], p=.052, Supplemental Figure 8A). When we included the participant with a protocol deviation (i.e., whose post-rTMS MRI was 12 days after rTMS instead of ≤ 7 days), the effect on DMN connectivity was reduced (t(9)=-0.82, 95% CI -1.97 to 0.32, p=.139, Supplemental Figure 8B).

**References**

1.	Tamminga CA, Ivleva EI, Keshavan MS, Pearlson GD, Clementz BA, Witte B, et al. Clinical phenotypes of psychosis in the Bipolar-Schizophrenia Network on Intermediate Phenotypes (B-SNIP). Am J Psychiatry. 2013;170(11):1263–74.

2.	First M, Spitzer R, Gibbon M, Williams J. Structured Clinical Interview for DSM-IV-TR Axis I Disorders, Research Version (SCID). New York: New York State Psychiatric Institute; 2002.

3.	Ashburner J, Friston KJ. Unified segmentation. Neuroimage. 2005;26(3):839–51.

4.	Friston KJ, Williams S, Howard R, Frackowiak RS, Turner R. Movement-related effects in fMRI time-series. Magn Reson Med. 1996;35(3):346–55.

5.	Behzadi Y, Restom K, Liau J, Liu TT. A component based noise correction method (CompCor) for BOLD and perfusion based fMRI. Neuroimage. 2007;37(1):90–101.

6.	Ashburner J. A fast diffeomorphic image registration algorithm. Neuroimage. 2007;38(1):95–113.

7.	Raichle ME. The restless brain. Brain Connect. 2011;1(1):3–12.

8.	First M, Williams J, Karg R, Spitzer R. Structured Clinical Interview for DSM-5—Research Version (SCID-5 for DSM-5, Research Version; SCID-5-RV). Arlington, VA,: American Psychiatric Association; 2015.

9.	Huang YZ, Edwards MJ, Rounis E, Bhatia KP, Rothwell JC. Theta burst stimulation of the human motor cortex. Neuron. 2005;45(2):201–6.

10.	Ward HB, Beermann A, Nawaz U, Halko MA, Janes AC, Moran LV, et al. Evidence for Schizophrenia-Specific Pathophysiology of Nicotine Dependence. Front Psychiatry. 2022;13:804055.

11.	Eldaief MC, Halko MA, Buckner RL, Pascual-Leone A. Transcranial magnetic stimulation modulates the brain's intrinsic activity in a frequency-dependent manner. Proc Natl Acad Sci U S A. 2011;108(52):21229–34.

12.	Yeo BT, Krienen FM, Sepulcre J, Sabuncu MR, Lashkari D, Hollinshead M, et al. The organization of the human cerebral cortex estimated by intrinsic functional connectivity. J Neurophysiol. 2011;106(3):1125–65.
